# Insecticide-treated bednet coverage in Africa 2006-2024: a spatiotemporal analysis of net ownership, use, age and type

**DOI:** 10.1101/2025.11.25.25341025

**Authors:** Eugene Tan, Mauricio van den Berg, Adam Saddler, Camilo Vargas, Daniel J. Weiss, Amelia Bertozzi-Villa, Samir Bhatt, Tasmin L. Symons, Peter W. Gething

**Affiliations:** Malaria Atlas Project, The Kids Research Institute Australia, Perth Children’s Hospital, Nedlands, WA, Australia; Institute for Disease Modeling, Gates Foundation, Seattle, WA, USA; School of Population Health, Curtin University, Bentley, WA, Australia; Department of Public Health, University of Copenhagen, Copenhagen, Denmark

**Keywords:** malaria, public health, interventions, epidemiology, mathematical modelling

## Abstract

Since their rapid adoption from the mid 2000s, insecticide-treated nets (ITNs) have been one of the most ubiquitous and impactful malaria interventions in Africa. Facing insecticide resistance and ever-increasing resource constraints, there is a growing need to revise previous modelling efforts of ITN coverage to better reflect the current epidemiological reality. Comprehensive estimates of ITN coverage must account for factors such as net type, net age, and user behaviours. By leveraging improved data availability, we propose a mathematically rigorous and interpretable ITN coverage model that extends previous national-level ITN models to the subnational level across 44 countries in Africa. The presented Multitype-ITN (MITN) model is used to construct longitudinal estimates of net demography by age and type from 2006 to 2024, and provide insights on potential sources of ITN distribution inefficiencies. Our findings show a significant increase in the uptake of next-generation ITNs, and thus reflect an increasingly diverse market. However, longitudinal coverage suggests stagnation in key metrics of net ownership such as nets-per-capita, access and use, alongside a downwards trend in utilisation – number of users per net. Coupled with observed stationary levels of use rate – proportion of people with access that also use nets. There are also early signs of interventions failing to meet the demands of increasing populations. Our results provide more nuanced metrics of ITN coverage that account for factors such as type and age, which may inform the design of future net allocation policies.

## 1 Introduction

Insecticide treated bednets (ITNs) have been the single most impactful tool in reducing malaria prevalence in Africa over the past twenty-five years, with approximately 2.5 billion bednets distributed to populations at risk across the continent over that time. ITN use has resulted in an estimated 68% of the observed reduction in infection prevalence [1–3] and a corresponding reducing in case incidence[3–8]. ITNs continue to play a critical role in malaria control, and the accurate tracking of ITN coverage is essential to monitor progress towards international coverage targets, enable evaluation of ITN impact on malaria transmission and burden, identify populations with inadequate protection, and understand the effectiveness of ITN distribution systems in reaching target populations [2, 9–11].

Information on ITN coverage comes from three main sources. First, nationwide community surveys capture detailed data on bednet ownership and use by households, providing rich intermittent snapshots of coverage across a given country. Second, operational data reported by national malaria programs (NMPs) detail the volume of ITNs distributed within a country (e.g., annual totals for national or subnational administrative units). Lastly, data from ITN manufacturers enumerate the volume of ITNs procured by countries over particular time periods. Each of these sources provide distinct and complementary information on the flow of ITNs through national distribution systems and into households, and the resulting coverage achieved.

While informative, simple characterisation of net coverage based on aggregated delivery and distribution reports does not capture the complex and dynamic nature of coverage and resulting epidemiological efficacy. Coverage metrics must account for a variety of factors such as net type, distribution equity, usage behaviours, and durability [2, 12–16]. Capturing these complexities is becoming increasingly important given the rapidly evolving landscape of new bednet products, growing insecticide resistance, and insecure malaria control financing [17] [18]) [19]).

A number of distinct catgories of ITNs can be defined. Early ITNs, now often termed ‘conventional’ insecticide treated nets (cITNs) predominated until around 2010 before begin phased out and replaced with long-lasting insecticide treated nets (LLINs), a washable and more durable product. More recently, ‘next generation’ LLINs have been developed with new classes of insecticide to counter growing insecticide resistance, with PBO based variants (e.g. G2, ROYAL Guard) are being increasingly deployed in the years following 2018 [19].

The modelling of ITN coverage has undergone several iterations with the earliest being the study by Noor et al. [20] and the spreadsheet-based NetCALC model presented by Killian et al. [21]. Early attempts at estimating ITN crop using historical data was proposed by Flaxman et al. where a compartmental “stock-and-flow” (SNF) model was used to track the flow of nets from manufacturer delivery to community distribution and finally removal (i.e. attrition) [22]. Bhatt et al. [2] substantially expanded the SNF model to include the use of more sophisticated attrition functions, missing data imputation, and spatiotemporal analysis. Bertozzi-Villa et al. [12] further refined the Bhatt framework and expanded its geospatial metrics to include nets-per-capita as well as access and use, hereby referenced as the Bhatt-BV model for brevity.

Since its development in 2021, The Bhatt-BV model has remained the state-of-the-art model for estimating ITN coverage. However, the diversification of the ITN market into multiple net types and greater interest in subnational level modelling now necessitates that the functionality of this approach is expanded. Differences in attrition and efficacy across net types means that quantifying and forecasting of the overall impact of ITNs must now account for net age and composition. Elements of the underlying compartmental model can also be enhanced to improve predictive performance, as described in by Tan et al. [23].

In this paper, we propose an updated multitype-ITN (MITN) model that extends the Bhatt-BV framework in the following ways (see Figure 1). (i) Data collection and model prediction is updated to reveal trends in ITN coverage at a national and continental level extending to the present. (ii) We leverage recent increases in the availability of sub-nationally resolved ITN distribution records, enabling a more granular understanding of coverage trends. (iii) We stratify modelling by ITN type and age, allowing the roll-out of next-generation ITNs to be tracked providing the first estimates of net demography and age dynamics. (iv) Finally, we revisit questions on systematic efficiencies in ITN distribution efforts and evaluate how this has evolved over time.

**Fig. 1.**
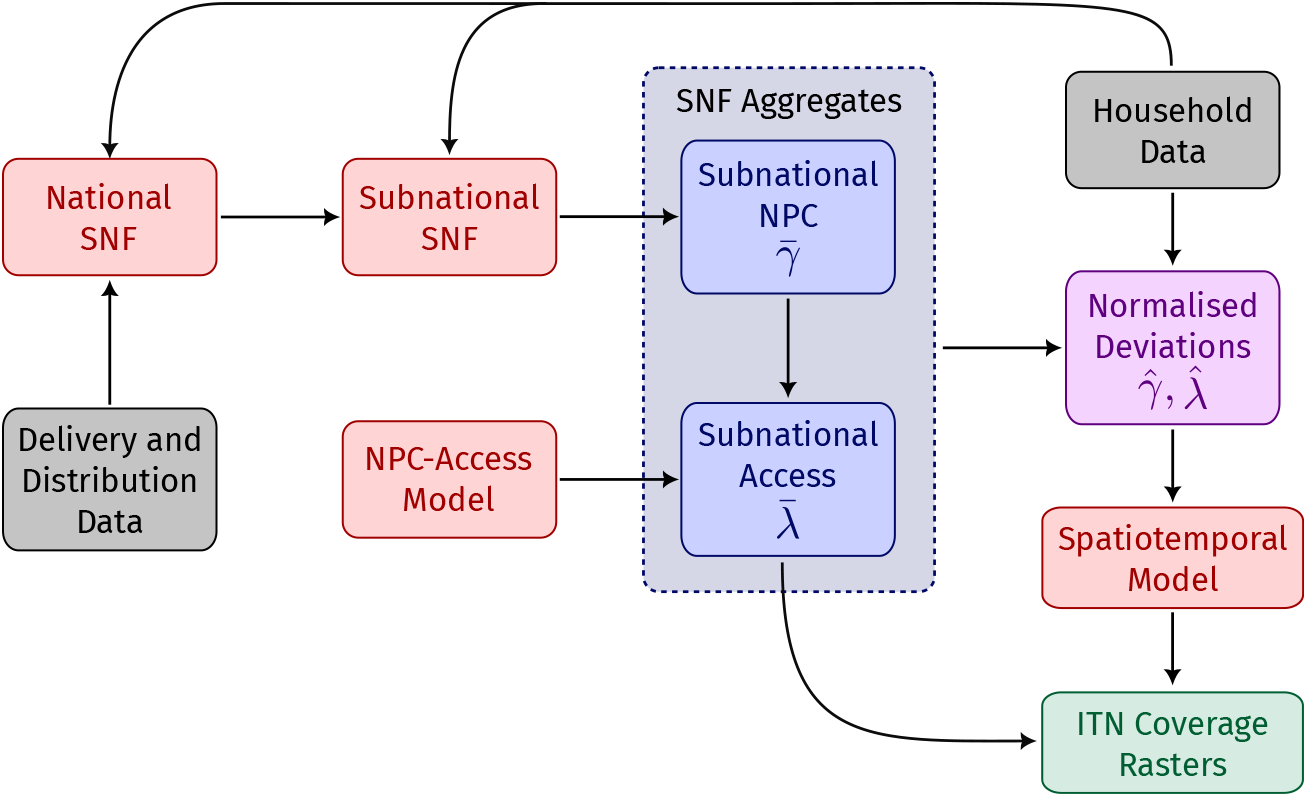
Schematic of the MITN model. Shown by color: model subcomponents (red), training data (grey), SNF outputs (blue) and final model outputs (green). Abbreviations: Stock-and-flow (SNF), nets-per-capita (NPC), insecticide-treated nets (ITN)

We consider six different ITN coverage metrics in our analyses. The first three are nets per capita (NPC) (*γ*), net access (*λ*) and net use (*ξ*). We retain the same definitions for these metrics for consistency with the existing gold standard Bhatt-BV model. NPC is defined as the ratio between net crop (Γ) (the total number of nets present in households) and population (*P*). Net access is the proportion of people who have access to a net in their household, where access is defined as “one net shared between two people”. Net use is the proportion of people in a population who use a net, where usage is indicated by a proxy that an individual has slept under a net in the night before a survey was conducted. We also study three other metrics: utilisation (*η*) is the estimated proportion of the net crop that are actually in use, use rate (*α*) is the proportion of people with access that also use a net. Finally, distribution factor (*ζ*) is the estimated proportion of underutilisation – *relative to the ideal case of K* = 2 *users per net* – that is attributed to suboptimal household net allocations. Notably, the distribution factor is highly nonlinear is used primarily to quantitatively describe the overall inefficiencies of net distribution programmes rather track longitudinal changes in ITN coverage.

For brevity, a summary of the model changes from the Bhatt-BV model is presented in the Methods section. An analytical approach to interpreting the dynamics and design of stock-and-flow models has also been discussed by Tan et al. [23]. Extended mathematical detail on the model is provided in the attached Supplementary Information.

## 2 Results

### 2.1 ITN Trends

#### 2.1.1 Population Coverage Metrics

After a long period of year-on-year increases, ITN coverage in Africa has plateaued since around 2017 (see Figure 2). Estimates for 2024 are that, on average, 48% (95%CI = 45-51%) of the population of malaria-endemic Africa had access to an ITN in their household, and 40% (95%CI = 35-46%) slept under an ITN. These metrics continue to fall well short of the WHO target of ‘universal’ coverage of 80% of the population protected [24]. Comparison of the year-on-year change in ITNs distributed in each country, versus the underlying growth in populations provides further context for the lack of progress - with distributed volumes largely offset, or even surpassed, by annual population growth in most years since 2017 (see Figure 7 in Supplementary Information). Specifically, this conclusion is based on the 5 year moving average of year-on-year change that smooths discontinuous perturbations due to mass campaigns, and tracks the macroscale availability of ITNs as a resource linked to standard economic metrics such as populations.

**Fig. 2.**
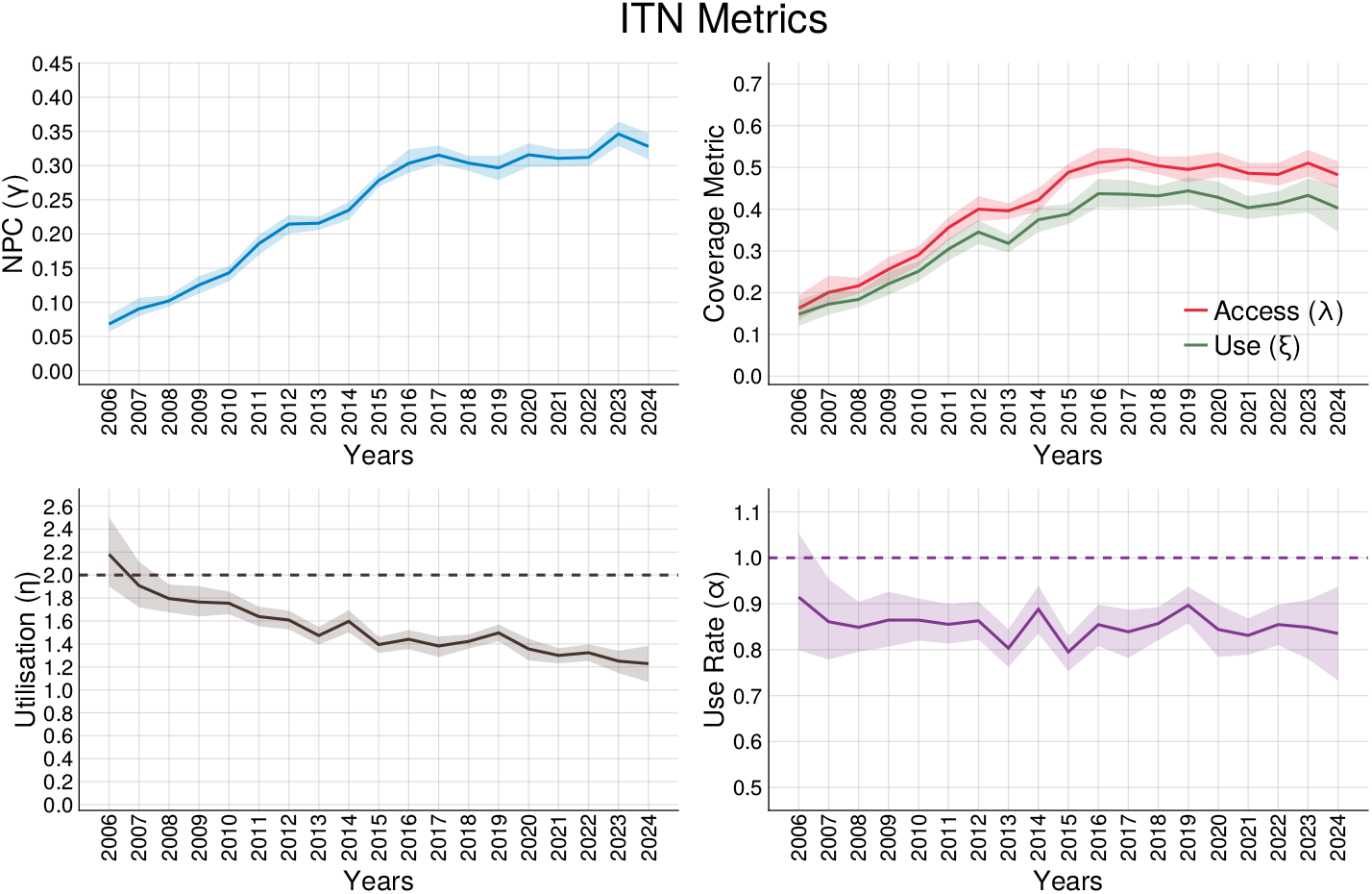
Weighted mean ITN coverage metrics aggregated across 44 countries in the African region. Shaded areas represent 95% credible interval. Nets per capita NPC (Γ) - the ratio between net crop (the total number of nets present in households) and population (*P*). Access (*λ*) - proportion of population who have access to a net in their household, where access is defined as “one net shared between two people”. Use (Ξ) - proportion of population who use a net in the night before a survey was conducted. Utilisation (*η*) - estimated proportion of the net crop that are actually in use. Use rate (*α*) proportion of population with access that also use a net.

Trends for individual countries are less straightforward to characterize, because of the periodic nature of mass ITN distribution campaigns and the resulting dynamic ebb and flow of coverage (see Figure 3). Nonetheless, when assessed over the medium-term, some countries display a broadly stable trend, with successive mass campaigns achieving similar levels of peak coverage (e.g. Uganda, Benin, Madagascar). Elsewhere, medium-term coverage is trending upwards, with recent years seeing new maxima achieved with large campaigns (e.g. DRC, Congo, CAR, Liberia, Niger). Conversely, countries including Kenya, and Senegal show evidence of a medium-term decrease in coverage from previous peaks, although this needs to be interpreted in the context of deliberate targeting of ITNs to subnational populations.

**Fig. 3.**
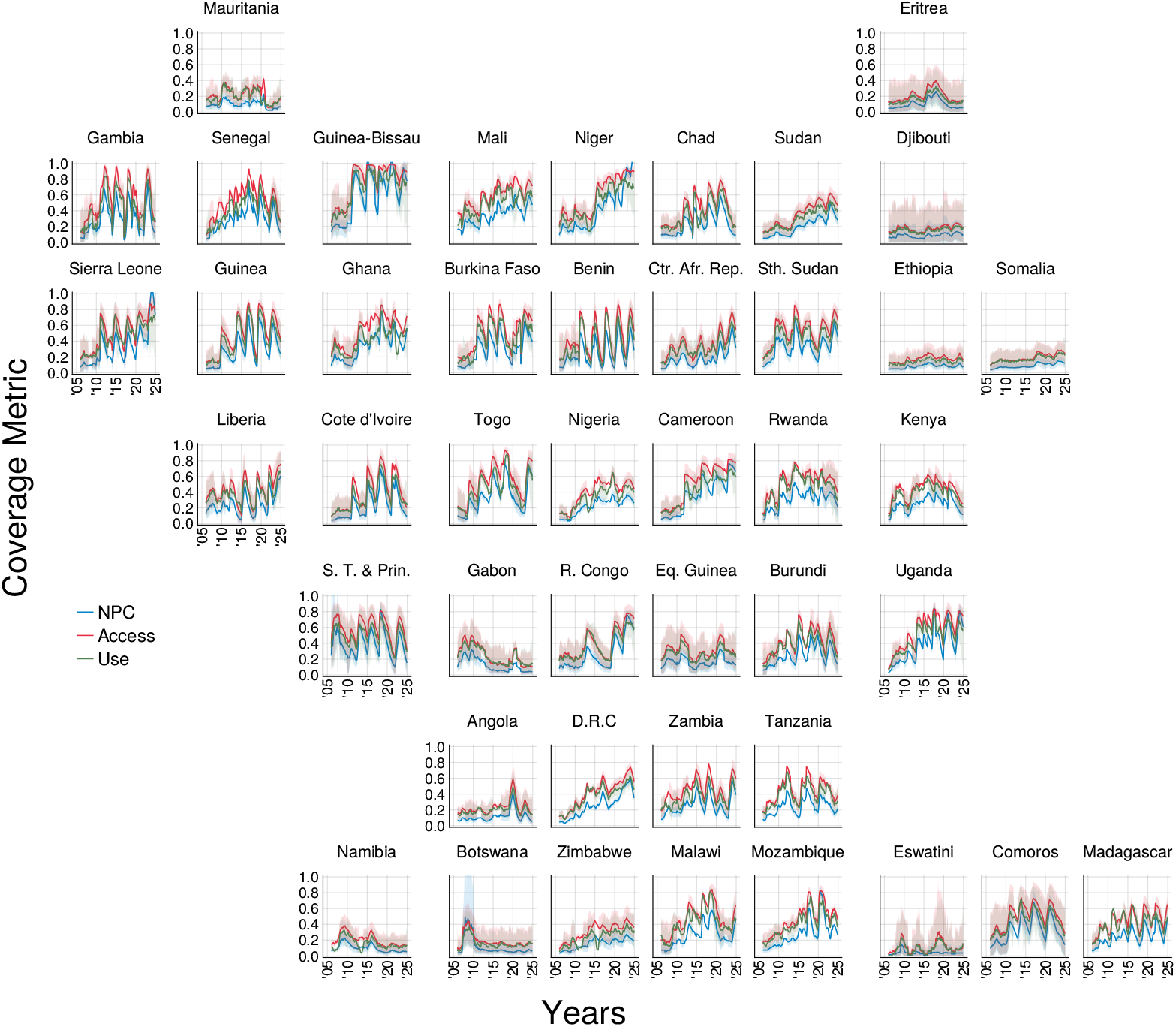
ITN coverage metrics for 44 modelled countries over the year period of 2000-2023. Shaded areas represent 95% credible interval.

#### 2.1.2 Net Demography and Supply

A key innovation of the MITN model is the capability to produce net demography estimates, enabling an understanding of not just the extent of coverage, but the type and age structure of nets being used as a key contributor to their likely physical integrity and insecticidal efficacy. The continent level aggregated estimate of net crop by age (see Figure 4, left panel) demonstrates how younger nets (less than 1 year old) make up a much smaller fraction of the continental total in 2024 (52%) shown in blue, than do old nets (10% are more than 2 years old) shown in orange-red.

**Fig. 4.**
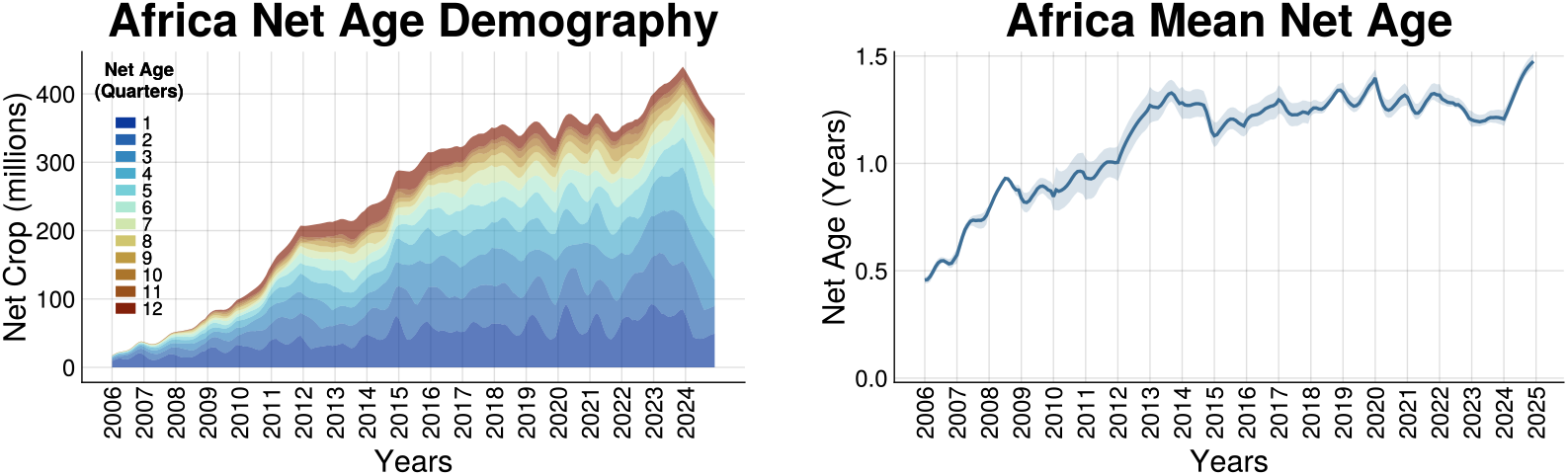
Continent level breakdown of net crop by age (left), and the mean net age (right).

Figure 4 (right panel) shows how the average age of nets in households has increased over time, with the era of rapid scale up prior to about 2015 characterised by lower average ages, and the subsequent period stabilising at a mean net age of around 1.25 years. This estimate broadly matches reported household net ages from DHS surveys where the mean net age for the period from 2015 to the present is approximately 1.48 years. The difference in reported and model estimates may be attributed uncertainties associated with household reports of estimated net age. Figure 5 (left panel) shows the various important transitions in net type over the past 20 years with the initial abrupt transition between cITNs and LLINs commencing around 2010, and the subsequent near-ubiquitous use of LLINs beginning to give way to the introduction of dual-active-ingredient (Dual-AI) and PBO nets from around 2018 (see Supplementary Information for country level estimates) with significant levels of distribution in next-generation of LLINs (PBO and Dual-AI) occurring in Central Sub-Saharn Africa. We estimate that these next-generation nets collectively account for around 34% of all nets in use in Africa at the outset of 2024, with that fraction set to rapidly grow. Figure 5 (right panel).

**Fig. 5.**
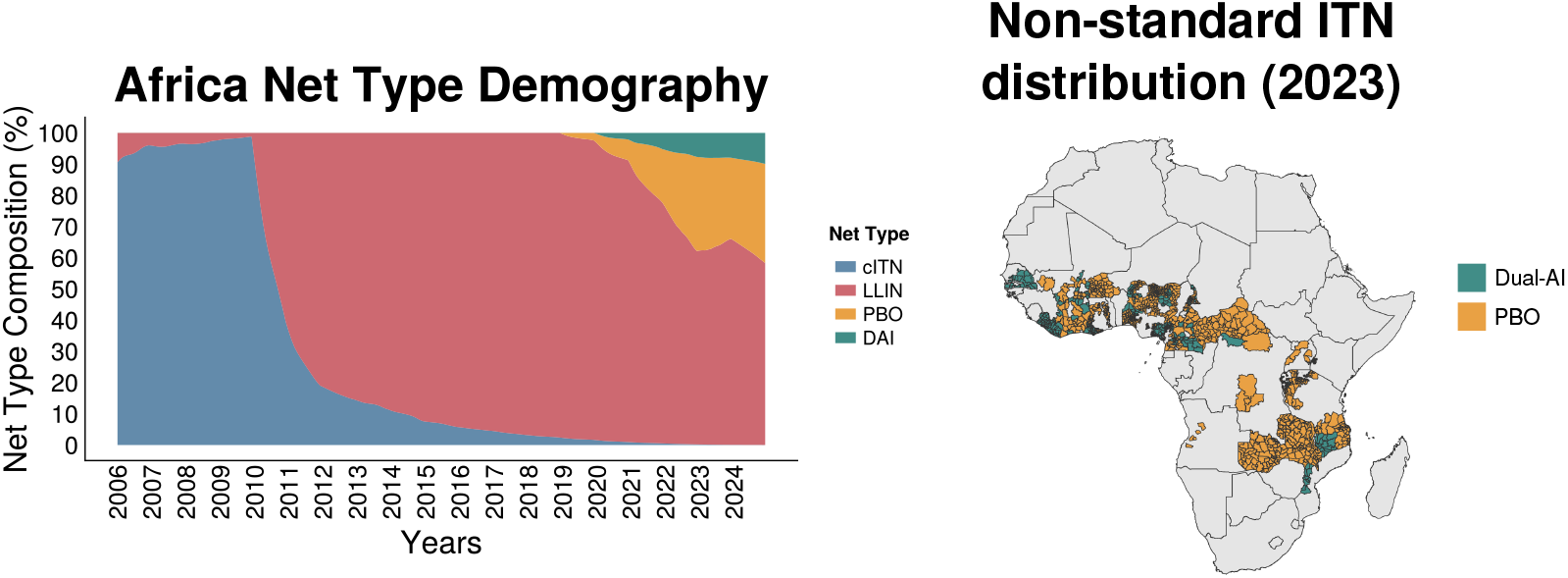
Composition of net crop market share by type (left), and reported primary net distributions of Admin 2 district level regions in 2023 for a selection of countries in Central Sub-Saharan Africa (right).

#### 2.1.3 Geospatial predictions

Figure 6 shows outputs from the geospatial model predicting NPC, access, use and mean net age at 5×5 km resolution for four different years across the study period. The maps for NPC, access and use portray the general pattern of increasing net provision and coverage between 2010 and 2020. Comparison of the 2020 and 2024 maps demonstrates the heterogeneity of trends across the continent, with different national and sub-national locations showing divergent trends over that period. The most recent coverage map (2024) demonstrates highly variable levels of contemporary coverage - partly reflecting that different countries are at different points within their cycle of mass campaigns. This is further showcased in the maps for mean net age - with those countries experiencing more recent campaigns having much younger nets, on average than those still reliant on a historical campaign distribution. The granularity of spatial patterns resolved in these maps varies considerably - with notably more resolved variation in those countries for which recent sub-national distribution data have been recently made available (e.g. Nigeria, Sudan, DRC).

**Fig. 6.**
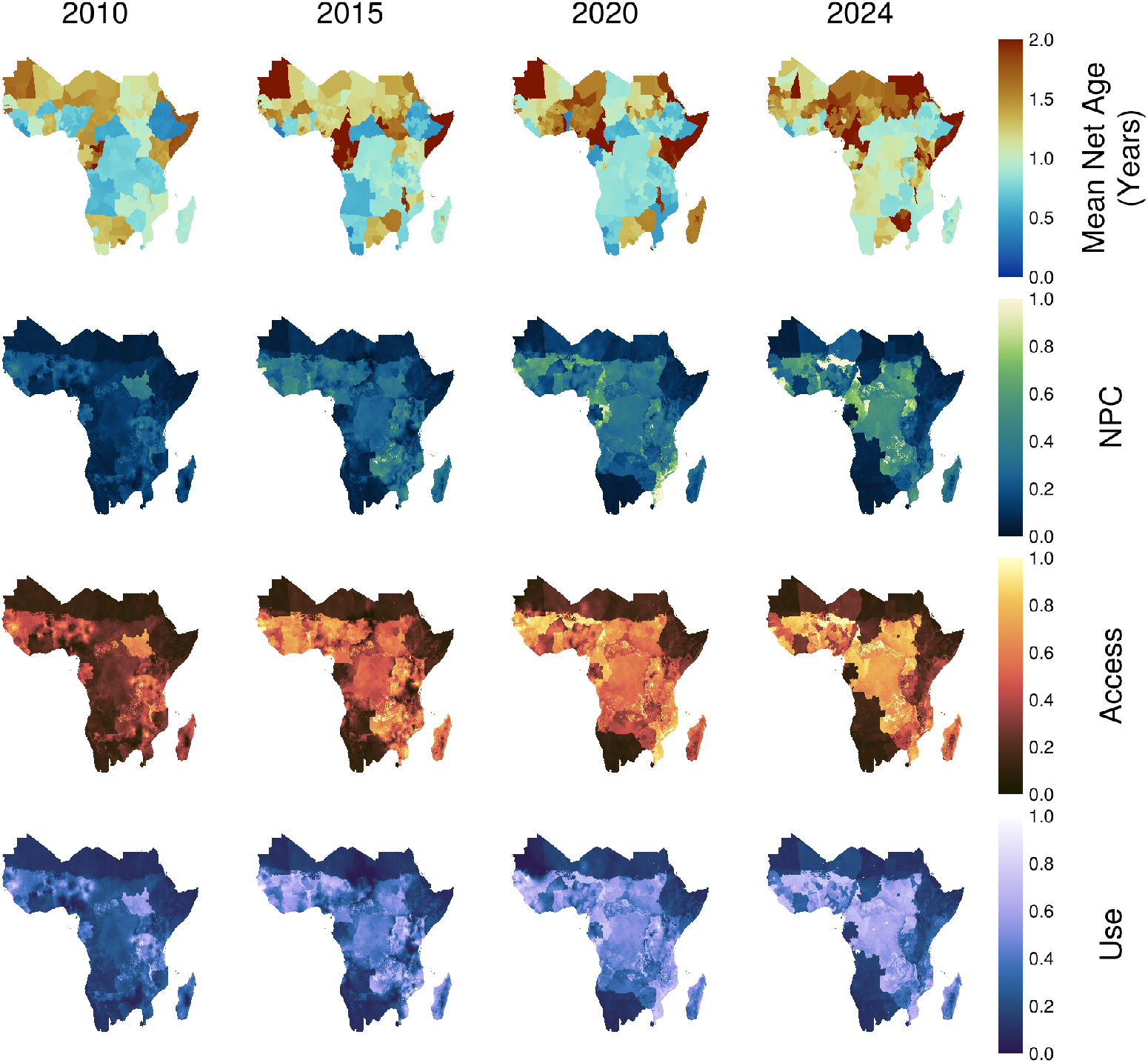
Maps of ITN coverage metrics for the years 2010, 2015, 2020 and 2023.

### 2.2 ITN Usage Behaviour Insights

#### 2.2.1 NPC - Access

One of the key findings presented by Bertozzi-Villa et al. was the nonlinear saturation of access with increasing NPC, preceded by a linear scaling region. Previous estimates indicated that for NPC values below 0.25, access and NPC scale linearly at a ratio that broadly aligns with the WHO recommendations of 1.8 people per net when calculating net allocations [25, 26]. Similar to findings by Bertozzi-Villa et al., at moderate to high NPC values, further increases in access behave nonlinearly and there are diminishing returns for increasing NPC [12]. However, we note that the NPC-access conversion model does not strictly enforce the condition that a net-ownership of zero coincides with zero access. To this end, we perform similar analyses to further investigate this inter-metric relationship. These comparisons are presented in Figure 7. Three levels of comparisons are conducted. The first identifies relationships in the outputs of the SNF component at different theoretical household age distributions (uniform and Poisson). The second relationship repeats this analysis on empirically calculated household size distributions taken from surveys. Finally, this comparison is extended to the full MITN model accounting for geographical heterogeneity.

**Fig. 7.**
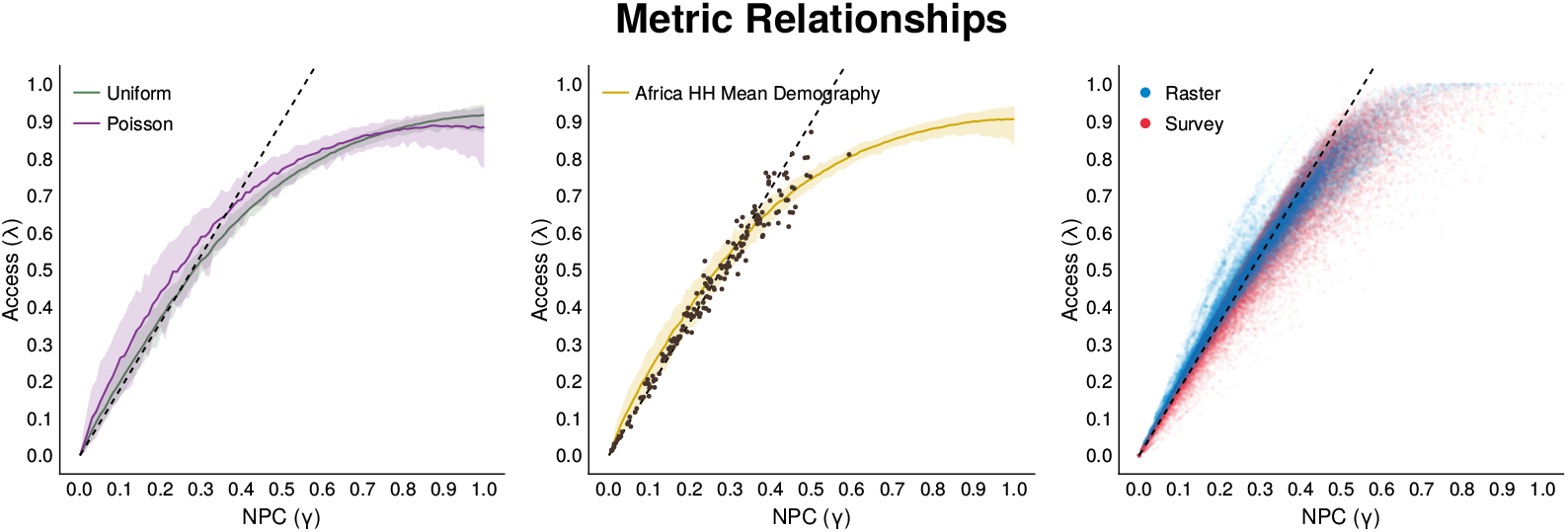
Relationships between NPC and access for different household size distributions. Simulated NPC-access using uniform and Poisson distributed household sizes (left), and empirical mean household demographic distribution case showing a good fit against survey-country observations shown as points (*N* = 175)(middle). Cluster level NPC and access showing saturation behaviour in both survey observations (*N* = 44853) and model estimates (right). Linear scaling lines shown represent a slope of 1.8 suggested by the WHO for determining net acquisition volumes.

For the first comparison, we find national values of NPC and access follow a mostly linear relationship with the scaling ratio of 1.8 providing a good fit for most of the data. However, nonlinear deviations from this trend are only seen to occur for NPC greater than 0.4. This onset is substantially higher than the previously estimated value of 0.25 [12]. We expand on the same analysis here by comparing the linear fit directly against observed data (shown as points), and also against theoretical model estimates (shown as lines).

To better understand the mediating factors for the presence of the linear region in the SNF model, estimates of NPC and access using various household size distributions evaluated at different NPC values were simulated. Three different household size distributions were compared: uniform *h* ~ *Unif* (0, 10), truncated Poisson *h* ~ *Poisson*(5), and an empirical distribution calculated from the aggregation of the entire household survey dataset across all countries in Africa (see Figure 7). We include the Uniform case to represent a control where household size distribution does not impact net access.

We find that in the current formulation of the SNF model, the distribution of household sizes are a key factor in determining the nonlinearity in observed scaling behaviours between NPC and access. Uniform household size distributions showed the clearest linear scaling region, whereas Poisson distributions revealed nonlinear scaling across almost the entirety of the NPC domain. Furthermore, in the case of a Poisson style household size distribution, access scales more efficiently at lower values of NPC. Simulations using empirical estimates of average African household size distributions show an intermediate behaviour between the former two. In all cases, there is significant overlap in the calculated credible intervals. However, it is worth noting that the means and size of credible intervals differ between each scenario. This finding suggests that proper care must be taken to account for the effect of household size distribution in the calculation of access and provides more support on the need to use a model that is more complex than a simple linear model as hinted by direct observation of the data. Extending similar analyses to spatially disaggregated cluster level predictions using the full MITN model, we find a strong match between predicted and observed values. Similar to the SNF comparisons, the onset of the nonlinear saturation regime occurs at approximately an NPC of 0.4.

#### 2.2.2 Utilisation and Distribution Factor

To better understand potential vulnerabilities and inefficiencies in the net coverage status of countries, we propose a second group of new ITN metrics: utilisation rate (*η*), distribution factor (*ζ*) and mean net age. The utilisation rate, defined as the ratio between use and NPC, is an approximate measure of the number of users per net owned (see Methods).

Whilst the utilisation rate is a rough descriptor of inefficiencies from reported distribution to use, it does not directly offer an explanation for the cause. We note two main mechanistic causes for underutilisation: (1) suboptimal tailoring for net distribution according to household size, and (2) non-use among households that have access to a net. The effect of the first also arises when studying the nonlinear scaling behaviour of the NPC-Access relationship, where increased net ownership does not necessarily translate to a proportional increase in access (see Figure 7). The second is described by the use rate, defined as the ratio between use and access. The distribution factor *ζ* tries to quantify the relative contribution of each cause to an observed utilisation rate,

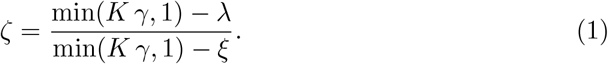

Broadly, the distribution factor *ζ* quantifies the proportion of underutilisation relative to an ideal case, that can be attributed to suboptimal allocation of nets based on household size. For our analyses, we define the ideal case as that of use rate equal to 1, and perfect linear scaling of access with nets per capita at a rate of *K* = 2 (i.e. based on the “two people per net” definition of access). For *ζ <* 1, the observed deviation from the ideal case is due to both suboptimal net allocation and reduced use rate with smaller values indicating a greater attribution to the latter. The case *ζ* = 1 corresponds to a perfect use rate and all underutilisation is due to suboptimal net allocation. Finally, *ζ >* 1 is the case where use exceeds access as users try to compensate for the lack of net access. A discussion on observed relationships is given in Section 2.2.3 and 2.2.4.

#### 2.2.3 Net Utilisation

Utilisation rates across the entire African continent appear to have a downwards trend with only an estimated 68% (95% CI = 53-69%) of distributed nets ending up in household usage if one assumes the ideal case of “2 users per net” (see Figure 2). This trend appears to be similar across smaller geographical scales with almost all countries showing either stagnating or downward trends in utilisation. Some notable exceptions are Kenya, Gabon and Chad sustaining modest but positive increases in utilisation rates (see Supplementary Information). Other examples such as Mauritania, Eritrea, Namibia and Botswana show volatile and erratic behaviour arising from poor data quality and availability.

Similar to net access, utilisation rate also has an interesting relationship with NPC. Both data and model predictions show a clear inverse relationship between utilisation and NPC (Figure 8). This suggests a tendency for nets to be underutilised when nets are abundant on a per-capita basis, which we discuss more below. The distribution factor (*ζ*) which describes the proportion of underutilisation attributed to sub-optimal ITN distribution is consistently below 1 (i.e. underutilised), but is most efficient at an NPC of 0.45.

**Fig. 8.**
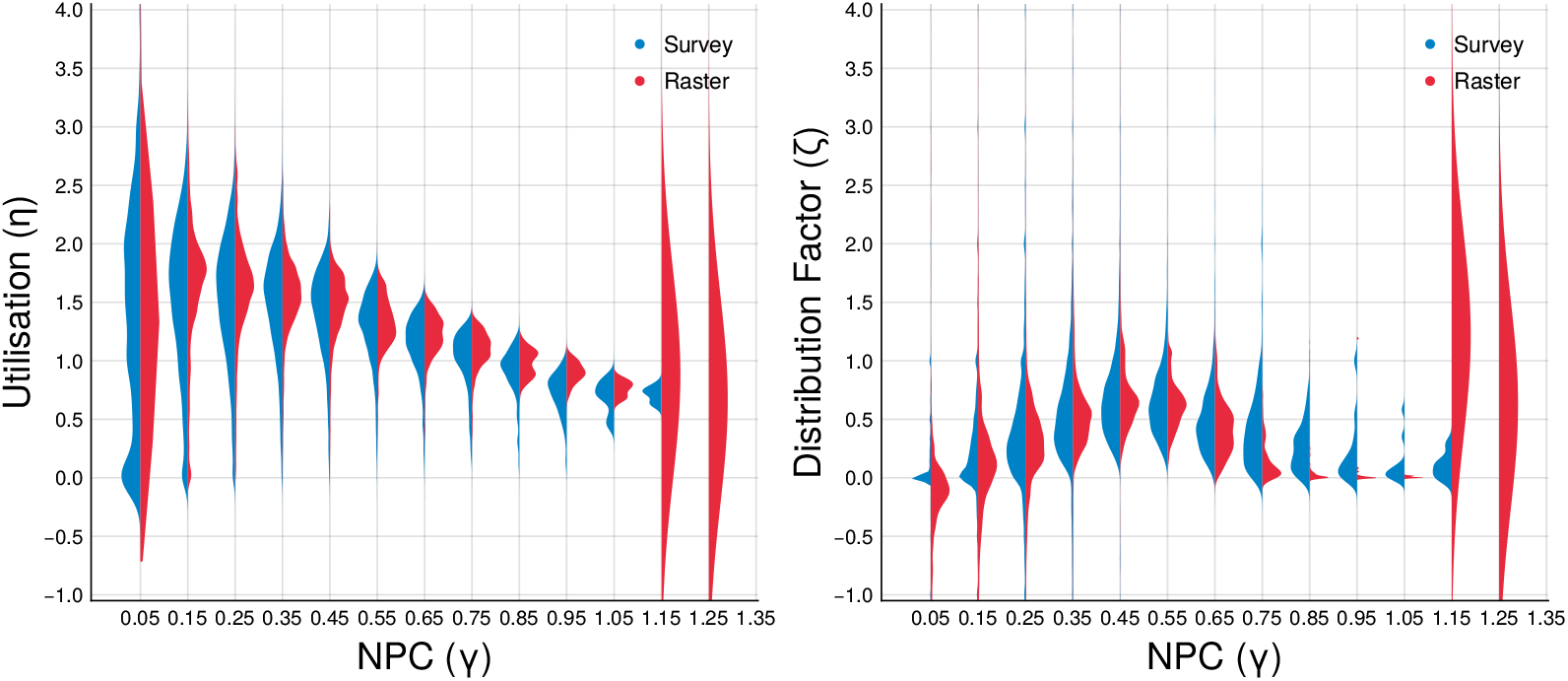
Nonlinear scaling relationships between utilisation (*ξ*) and distribution factor (*ζ*) and NPC (*γ*). Distributions for NPC bins of width 0.1 with observed values (blue) and model raster predictions (red). Large inaccuracies occur for extremely high observed and modeled NPC values as they lie well outside the domain captured by a bulk of the training data.

#### 2.2.4 Causes for ITN Inefficiencies

To investigate the potential sources of underutilisation, we observe that distribution factors in both survey and raster sources show a positive trend for increasing NPC until a critical value of approximately *γ* = 0.45. However, as NPC continues to increase, this saturates the household net access, *ζ* decreases and a larger proportion of underutilisation is attributed to a compromised use rate. Despite this trend, we find that all clusters with high NPC have *ζ >* 0 indicating the suboptimal net allocation across different household sizes remains a significant contributor to decreased net utilisation. Cases where *ζ <* 0 correspond to access and use scaling more efficiently than the reference *K* = 2 scenario, and usually are the result of numerical artifacts where NPC (*γ*) is small.

The observed underutilisation in nets can be separated into three main regimes by plotting Δ*λ* = min(*Kγ*, 1) − *λ* and Δ*ξ* = min(*Kγ*, 1) − *ξ*, which are the deviation of access and use from the ideal utilisation case (i.e. perfect linear scaling of NPC-Access, and maximum use rate). (see Figure 9). We find that an overwhelming number of observations indicate an underutilisation of nets with an increased attribution to suboptimal net allocation practices as NPC increases. Whilst there are cases where overcompensation in net use in both survey observation and models, and potential over utilisation above the expected ideal in model predictions, these cases are rare. Thus we conclude that suboptimal net allocation practices remain a challenge that needs to be addressed to tackle inefficiencies in ITN coverage initiatives.

**Fig. 9.**
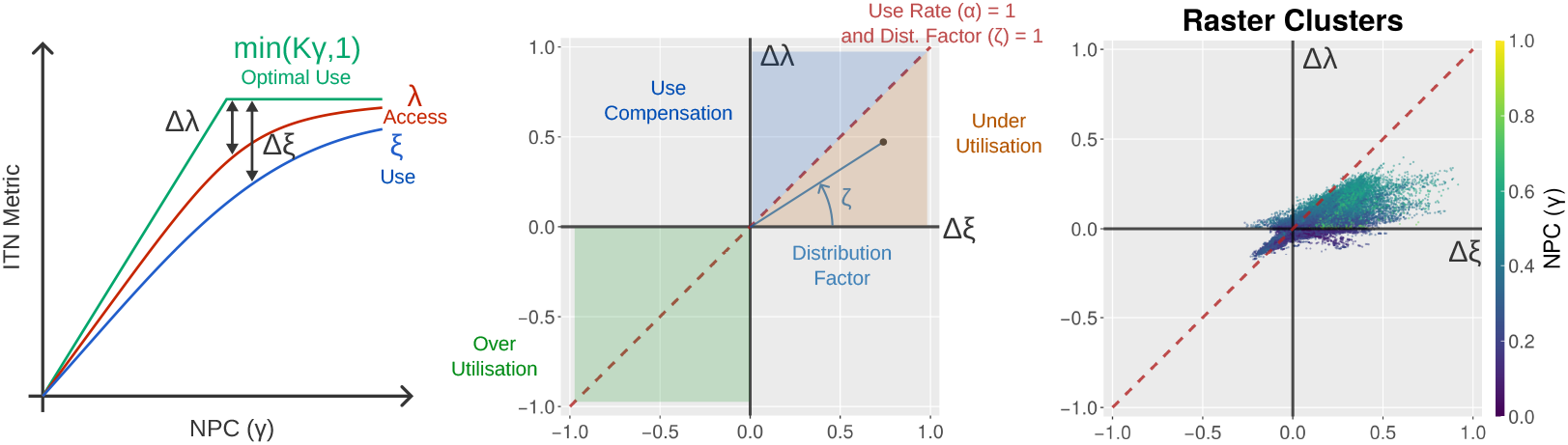
Plots of deviation in access and use from the ideal case (left). An interpretation guide with different regimes shown in the top row. The slope of the line from the origin to an observed point is equivalent to *ζ* (middle). Model outputs of the same plot (right).

## 3 Discussion

This paper presents recent innovations and extensions of the widely used Bhatt-BV model first proposed in 2021. In the four years since that last iteration of our ITN coverage models, the epidemiological and economic landscape of malaria has changed dramatically, and we have updated and extended the framework to address these new challenges and generate up-to-date insights into ITN coverage and system efficiency.

The proposed multitype-ITN (MITN) model includes numerous structural and mathematical changes to the previous framework. In addition to providing improved data fits and mathematical interpretability, the MITN model also includes enhancements in multiple key technical capabilities. The most notable of these include inference of ITN coverage at greater temporal resolutions from quarterly to monthly, and the capability to simultaneously model multiple net types. These features have been chosen to enhance the data parsimony and flexibility of the models as countries begin to move into a resource constrained a world with a more diverse net market.

We demonstrate the utility of the MITN model to provide updated insights on global trends of ITN coverage and user behaviour. In terms of the suite of traditional ITN metrics, our findings reveal a continuation of the prevailing narrative that the availability and uptake of ITNs has stagnated from 2018 Aligned with this, much of the African region still falls short of the WHO recommended threshold for universal ITN coverage. We also reveal that, for the first time since the start of widespread ITN usage, the growth in net availability has been outpaced by population growth across the African region.

We again find evidence that incremental increases in access require progressively larger increase in net supply once a certain threshold of access is reached - i.e. demonstrating diminishing returns. While our earlier work identified this threshold at an NPC of around 0.25 [12], this updated analysis concludes a higher level of around 0.4. Furthermore, we demonstrate that household size demography is a potential mediator for the prominence of the observed nonlinearity.

In addition to the NPC-Access relationship, we estimate the utilisation rate of nets across the African region. Across Africa, utilisation rates of ITNs have steadily decreased in recent years with similar trends occurring across a majority of countries. The relative impact of suboptimal net allocation policies and user adherence reveals that the former remains the most dominant factor of net underutilisation. As the prevalence of nets increase, attributions slowly shift to non-ideal use rates resulting from poor user adherence. However, this effect is relatively small with most countries not achieving a sufficiently high NPC for this to be a factor. In both observed data and model predictions, there is an observed inverse relationship between NPC and utilisation for NPC values greater than 0.35, which further supports the hypothesis of distribution inefficiency. Nevertheless, it is important to note that these relationships describe observed macroscale behaviours at specific levels of net ownership (NPC), and net retention and attrition still remains the biggest factor in determining changes in NPC.

Our analysis provides the first systematic, continent-wide estimates of net demography by type and age. Consistent with increasing diversity of net types in the market, current trends demonstrate the progressive uptake of next generation ITNs that may be more efficacious in areas with insecticidal resistance. Our analysis of net age demonstrates how many nets currently in use are at least two years old - and age at which physical and insecticidal integrity may be compromised [17]. Further work is needed o better quantify the effects of age on net bioefficacy, reliability, and performance in deployment. Coupled with the ability to estimate net demography, the MITN model presents an avenue of potential research where effective ITN coverage can be used to better track and describe the impact of ITN intervention programmes, accounting for nuances in ageing and declining net quality.

## 4 Methods

### 4.1 Data

The entire MITN model is calibrated against a collection of annual deliveries and distribution data, population data and household surveys spanning the period 2000-2023. Manufacturer LLIN delivery reports were obtained from the Alliance for Malaria Prevention’s (AMP) Net Mapping Project and compiles data from various sources including the Global Fund, the President’s Malaria Initiative (PMI), UNICEF, the Against Malaria Foundation (AMF), World Bank, UNITAID and several others. National level annual net distribution data were compiled from various sources including National Malaria Control Programs (NMCPs) provided by WHO, African Leaders Malaria Alliance (ALMA) and PMI, and contains estimated of distributed volumes classified by net type. Household surveys were collated from publicly available DHS and MICS datasets, with geolocation data available from the former. For countries with no household data, ITN metrics data points were estimated from available survey reports summaries. Population data was taken from the Institute for Health Metrics and Evaluation (IHME). A detailed breakdown of delivery and distribution datasets is provided in the Supplementary Information.

### 4.2 Model Assumptions

Several assumptions are required in order to provide sufficient structure to the ITN modelling problem such that calibration can be done with existing available data. The list of assumptions for each model component are given as follows.

Assumptions for the national and subnational net crop SNF components:

- Prior to 2010, distributions of nets consist of a mixture of cITNs and LLINs whose relationship adheres to a constant ratio unique to each country given by *α*_*LLIN*_
- No cITN distributions are made after 2010.
- Redistribution strategies *ϕ* are static between each year, but are unique to each country.
- Net deliveries arrive in full at the start of each calendar year and immediately add to the country’s net inventory

Assumptions for the NPC-Access conversion model

- The NPC-Access conversion is global and entirely determined by the household size distribution and net crop
- During prediction, household size distributions for each country are static and taken as the historical average across all available surveys for that country

Assumptions for the spatiotemporal disaggregation model

- Deviation metrics can be sufficiently explained using covariate indices

### 4.3 MITN Model Components

#### 4.3.1 SNF Net Crop Model

The SNF model for net crop consists of a compartmental model with extended gating mechanisms represented as a collection of difference equations. Analytical simplifications conducted by Tan et al. [23] show that such a model can be decomposed into either a linear time invariant system, or non-autonomous dynamical system. They also provide recommended methods for uncertainty inflation, model fitting and temporal disaggregation with numerical results on convergence properties.

The equations for the gating mechanism used to track excess stock, and reconcile delivery with distribution data is given by:

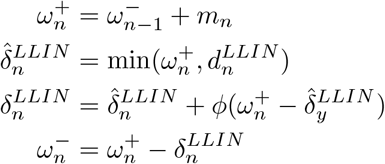

subject to the initial condition,

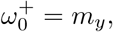

where variables are defined as follows:

- *m*_*n*_ : Reported number of LLINs delivered at the start of year *n*
- 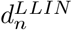 : Reported number of LLINs distributed at the start of year *n*
- 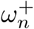 : Number of LLINs in the national stock at the start of year *n*
- 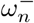 : Number of LLINs in the national stock at the end of year *n*
- 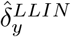 : Number of LLINs distributed excluding additional redistribution of current excess net stock
- 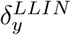 : Number of actual LLINs distributed during the entirety of year *n*
- *ϕ*: Unique redistribution constant for each country.

Because the number of cITNs distributed in any given year is unknown, a regressed conversion parameter is used to account for their potential distribution,

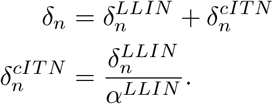

Annual inferred distribution time series of type *i* given by 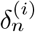 is temporally disaggregated using learned constant parameters,

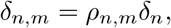

where *ρ*_*n,m*_ is the proportion of nets in the *n*^th^ year that are distributed in the *m*^th^ month of that year, and is subject to the constraint,

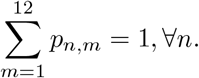

The validity of the temporal disaggregation approach assumes the spatial homogeneity in sampling of households between each month. This is checked using the Jensen-Shannon Divergence (JSD) criteria described in [23].

The natural attrition (removal) of nets is represented using a two parameter sigmoidal decay function with compact support as suggested by [21],

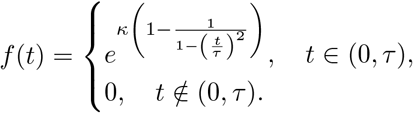

The above chosen form for the attrition function *f* (*t*) differs from the Bhatt-BV model [12] by using a more flexible two parameter form instead of a single parameter. A discussion on its approriateness is discussed in [23] alongside other potential functions.

Output timeseries of net crop from running the national SNF model 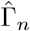 is calibrated against netcrop estimates calculated using household aggregated estimates of net crop per capita (NPC) multiplied by the population in each region (i.e. Γ_*n*_ = *γ*_*n*_*P*_*n*_). Volatility between monthly NPC national aggregate estimates are accounted for using a moving average filter and uncertainty inflations methods (see Tan et al. [23]). The resulting statistical regression is thus given by,

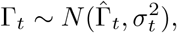

where *t* represents the month index starting from the first year, and *σ*_*t*_ is a calculated inflated standard error (see Supplementary Information and Tan et al.[23]).

Fitting of the SNF model is done using iterative expecation-maximisation algorithm that alternates between fitting point estimates of temporal disaggregation ratios *p*_*n,m*_, and random variable posterior estimates of attrition *τ, κ* and redistribution parameters *ϕ*. For the former, a stochastic gradient descent algorithm is used to optimise the DIC according to the following loss function,

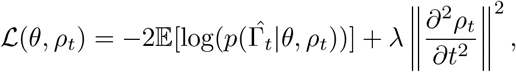

where the second term is a Laplace regulariser that penalises discontinuities and prevents overfitting, and *θ* represent all other learned model parameters (e.g. net attrition, redistribution etc.) A standard Metropolis-Hastings MCMC algorithm is used to calculate Bayesian posterior samples for remaining parameters *θ*. The algorithm is self-terminating when the estimates of ℒ fails to increase between each iteration cycle.

The MITN model requires two different rounds of attrition parameter estimation, one each for the national and subnational estimates. Attrition parameters *τ*_*nat*_, *κ*_*nat*_ for the national SNF parameters are first fitted to get an initial estimate. A second round of fitting is done using subnational distribution and household survey data (alongside their corresponding uncertainty inflation values) to estimate *τ*_*subnat,j*_, *κ*_*subnat,j*_ the attrition parameters for subnational region *j*. Here, national parameters *τ*_*nat*_, *κ*_*nat*_ are used to define prior distributions from which to compute Bayesian estimates. Further details are provided in the Supplementary Information.

#### 4.3.2 Access Model

The NPC (*γ*) - access (*λ*) conversion model is centred around the estimation of the joint distribution *H*(*n, h*) where *n* is the number of nets owned by a household *n* and *h* is the household size. Similar to the Bhatt-BV model, a zero truncated Poisson (ZTP) model is used [2, 27] with two intermediate model variables is used to describe the joint distribution,

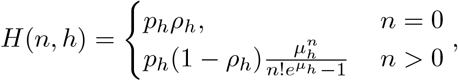

The intermediate model variables are *ρ*_*h*_ representing the proportion of households of size *h* with no nets, and *µ*_*h*_ the mean number of nets owned by a household of size *h*. Each variable is represented by a flexible general statistical function. The model for *ρ*_*h*_ is given by,

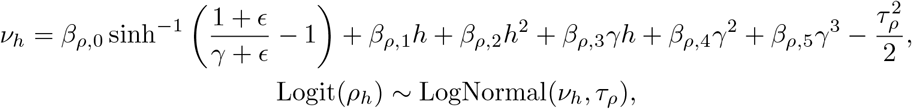

where *β*_*ρ,i*_ and *τ*_*ρ*_ are free parameters. The inverse hyperbolic sine term is included to ensure that *λ* → 0 for *γ* → 0, a constraint that was not upheld in previous models [2, 12].

A similarly general function is used to model *µ*_*h*_,

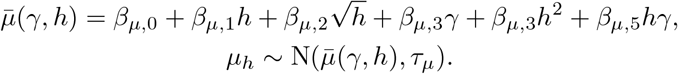

This formulation simplifies the approach from [2, 12] by using a single unified nonlinear model rather than a stratified set of linear models unique to each value *h*. This significantly reduces the number of parameters in the model, and allows for prediction on values of *h* that were not originally in the training domain.

Population level access *λ* is calculated using the joint distribution using the following expression based on the definition of access being “one net between two people per household”,

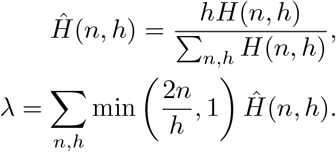

A global NPC-Access conversion model is fitted using the entirety of the household survey dataset. As the above model formulations are differentiable, a NUTS MCMC [28] sampler was used to optimise for convergence and computation time.

#### 4.3.3 Spatiotemporal Disaggregation Model

Spatiotemporal disaggregation of SNF subnational aggregates into high resolution rasters of 5×5 km pixels is done by constructing models of normalised local deviations as a function of a collection of time-varying spatial covariates. This general approach is similar to that used by Bertozzi-Villa et al. in the Bhatt-BV model. However, the original approach of defining deviations as an arithmetic gap (NPC gap, access gap and use gap) is not mathematically proper as they fail to account for the dependence between aggregate values and local deviation size. The usage of arithmetic gap also does not restrict the resulting estimated local ITN coverage metrics to the correct domain. To correct this, three different normalised local deviations are used: NPC deviation 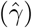, deployment rate 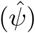 and use-access deviation 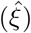. For notation, bars represent SNF subnational aggregate estimates, and *x* are spatial locations.

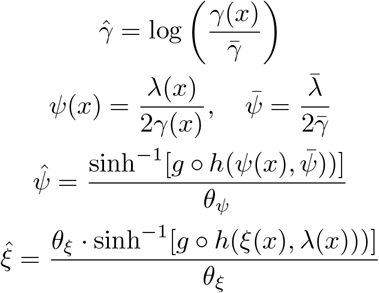

where functions *g, h* are transformations used to ensure the correct constraints for domains are adhered to (i.e. *γ*(*x*) ∈ [0, ∞), and *λ*(*x*), *ξ*(*x*) ∈ [0, 1]). These are defined as follows,

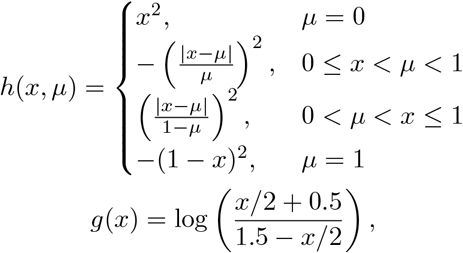

where *h* is an asymmetric function that maps the deviation of an observation *x* ∈ [0, 1] from a reference point *µ* ∈ [0, 1], and *g* is a logit function with a transformation applied such that no special bias is given to observed values of *x* = 0.5.

An inverse hyperbolic sine transformation is used in order to allow for a better conditioning of the regression problem. This transformation is subsequently scaled with a constant *θ*, chosen to maximise the concentrated log-likelihood with respect to the observed data [29].

Local deviations calculated from observed household values are used to train three different SPDE spatiotemporal models, which is regressed against a set of static, annual and monthly resolution covariates 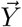 and random spatiotemporal field *F*. NPC deviation and deployment rate are regressed at an annual resolution, but use deviation is regressed at monthly rate to account for potential seasonality in usage. The general statistical model is thus given as

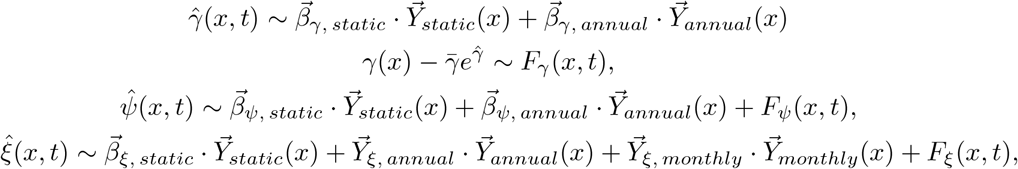

where the spatial component of the random field *F* is represented as Gaussian process with Matérn covariance, and the temporal component is a first order autoregressive process. Fitting is done using the R-INLA package [30].

## Supplementary information

This paper contains accompanying supplementary information. Please refer to attached documents.

## Acknowledgements

This work was supported, in whole or in part, by the Gates Foundation [INV-055192]. The conclusions and opinions expressed in this work are those of the author(s) alone and shall not be attributed to the Foundation. Under the grant conditions of the Foundation, a Creative Commons Attribution 4.0 License has already been assigned to the Author Accepted Manuscript version that might arise from this submission. This work also includes funding support from the Australian Government, National Health and Medical Research Council (Award No: GNT2025280).

## Declarations

### Data Availability

The household-level survey data used in this analysis is publicly available from the DHS (https://dhsprogram.com/) and MICS (https://mics.unicef.org/) websites. The national-level-aggregated survey data were gleaned from reports available at the MIS website (https://www.malariasurveys.org/). Data on manufacturer delivery of nets are available from the AMP Net Mapping Project (https://allianceformalariaprevention.com/working-groups/net-mapping/). Data on NMCP distribution of nets were provided by WHO.

### Code Availability

A repository containing related model code is publicly available on the repository. (https://github.com/eugenetkj98/MITN-Public). A compilation of model numerical outputs with high resolution can be provided at request.

