## Supplementary Information for "Insecticide-treated bednet coverage in Africa 2006-2024: a spatiotemporal analysis of net ownership, use, age and type"

#### 1 MITN Model Overview

Similar to the gold standard Bhatt-BV model [1], the proposed MITN model reconciles various sources of ITN data (manufacturer net delivery, community distribution and household survey data) to provide estimates of coverage metrics such as nets per capita (NPC), access and use. The MITN model builds upon the general architecture of the Bhatt-BV model and consists of 4 model components (see Figure 1).

First, a national SNF compartmental model is trained to reconcile delivery, distribution and household survey data to provide baseline national aggregate estimates of attrition parameters and net crop. These national estimates are further refined using a simplified subnational variant of the same model to account for regional heterogeneity in net coverage and attrition behaviour. Independent of the SNF models, a conversion model between NPC and Access is trained, the outputs of which are combined with subnational SNF outputs to provide subnational aggregate estimates of net access.

Subnational aggregate estimates are subsequently used to normalise geolocated household survey entries of net ownership (NPC), access and use and produce local deviation metrics. A spatiotemporal model is then constructed to regress local deviation metrics against a collection of a plausible covariates. Once trained, sampled deviation rasters are fed back into subnational aggregate estimates to produce interpolated high resolution global rasters of ITN coverage metrics.

Unlike previous iterations of ITN coverage models, the MITN model is designed to accommodate an arbitrary number of net types. This capability is particularly valuable as type diversity in the the market share of nets continue to increase. However, whilst the model does allow for posterior parameters estimates for each net type, the dearth of household survey data leads and recency of next-generation ITN adoption leads to an identifiability problem when accounting for multiple net types. This identifiability is not restricted to only next-generation ITNs, and in poor data cases, also impact the identifiability of attrition parameters between cITNs and LLINs (see SI Section 6).

The impact of poor data is heterogeneous across countries, with some notable countries such as Ghana, Nigeria and Mali leading in data quality and availability. Nevertheless, the MITN model provides the modelling infrastructure for such an exercise when more future data becomes readily available.

Recognising this data insufficiency, our analyses assume all forms of next generation ITNs considered (PBO, Dual AI) inherit the same attrition characteristics as the current first generation LLIN, which retains a majority of the net crop market share. Furthermore, due to the lack of accurate distribution data by type in the years prior to 2010 during which cITNs were still actively deployed, we assume almost all nets distributed prior to 2010 are cITNs. A small fitted conversion factor is included to account for the potential small proportion of LLINs that may have been distributed during this time.

#### 2 Data Sources

Three main datasets are used to construct ITN coverage models, which themselves are compiled from a collection of publicly available data sources. These datasets or annual net delivery data, annual net distribution data and household survey data of net use. Net delivery refers to the shipment of nets from manufacturers to national programmes and other other distributing bodies. Net distribution refers to the provision of nets from distribution bodies to members of the community. The datasets used in the analyses were inherited from those of the Bhatt-BV model [1] with data from subsequent years (2021-2004) appended from the same base sources and pre-processing. We refer to [1] for further technical details on data processing.

Data and analyses for the study was conducted for the period of 2000-2024. LLIN delivery data were primarily extracted from the AMP Net Mapping Project (https://allianceformalariaprevention.com/working-groups/net-mapping/) with additional aggregations from various reports sources from National Malaria Control Programs (NMCP), African Leaders Malaria Alliance (ALMA), and President's Malaria Initiative Operational Plans (PMI

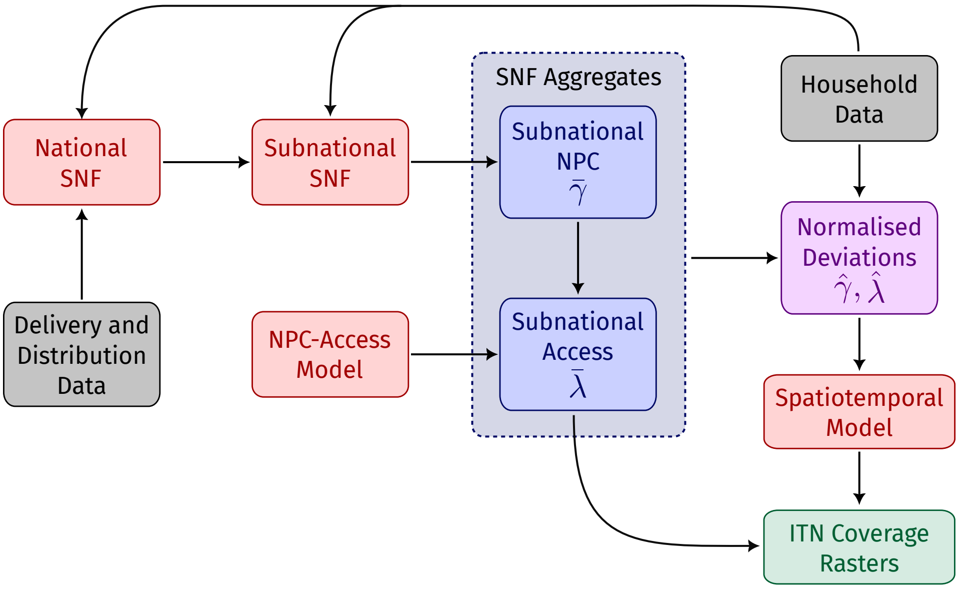


Figure 1: Schematic of the MITN model. Shown by color: model subcomponents (red), training data (grey), SNF outputs (blue) and final model outputs (green). Subnational components (blue) and normalised deviations (magenta) constitute the main additions of the MITN model.

MOP). Net distribution time series were primarily provided from WHO sources, with additional input from the National Malaria Control Programs (NMCP), ALMA and PMI. This dataset contains similar source data with those used by Bertozzi-Villa [1] with additional detail regarding net type provided by WHO. Reported numbers and subsequent estimates provide a time series of national net inventory and distribution given at an annual temporal resolution.

The household-level survey data used in this analysis is publicly available from the DHS (`https://dhsprogram.com/`), and MICS (`https://mics.unicef.org/`) websites. The national-level-aggregated survey data were gleaned from reports available at the MIS website (`https://www.malariasurveys.org/`. Data on manufacturer delivery of nets are available from the. A total of 189 household surveys across 41 countries were aggregated from multiple sources (MICS, MIS and DHS) and included values of number of cITNs, number of LLINs (type agnostic), net age and net use in the night prior to the survey. Surveys were further disaggregated by survey month and post-processed to provide 921 country-month estimates.

#### 2.1 Delivery and Distribution Data

##### 2.1.1 National Data

Manufacturer delivery data is only available up to a national resolution, with most countries having complete annual delivery time series from 2004 onwards. Data also only contains values for LLIN deliveries, with no cITN available. ITN distribution data is provided from WHO sources and contains type information in the years following 2020 where the distribution of next-generation ITNs occur. As type information prior to this point is not available, we assume that any reported net distributions that occur prior to 2010 are predominantly cITNs. A summary of available delivery and distribution data is shown in Table 1.

To account for potential inaccuracies in reported distribution data, the SNF model utilises delivery data to track a 'virtual' inventory of ITNs within a country in any given year, which is subsequently used to place an plausible upper bound of ITN distributions. Due to the lack of cITN delivery data, distributions of cITNs (i.e. prior to 2010) are not artificially bounded. Furthermore, a statistical conversion factor between LLIN and cITN distributions is applied as part of the national level SNF model for the period from 2004-2010 to allow for potentially small amounts of LLINs to be distributed if reported deliveries are non-zero.

Table 1: Data availability summary for national LLIN deliveries and ITN distribution.

| ISO | Country | LLIN Delivery Starting Year | ITN Distribution Data Years |
|---|---|---|---|
| AGO | Angola | 2004 | 2006-2022 |
| BDI | Burundi | 2004 | 2001-2024 |
| BEN | Benin | 2004 | 2006-2024 |
| BFA | Burkina Faso | 2004 | 2005-2024 |
| BWA | Botswana | 2006 | 2010-2012, 2015-2020 |
| CAF | Central African Republic | 2005 | 2001-2004, 2006-2010, 2012-2024 |
| CIV | Côte d'Ivoire | 2006 | 2003-2004, 2005, 2008-2011, 2013-2024 |
| CMR | Cameroon | 2004 | 2005-2024 |
| COD | Democratic Republic of the Congo | 2004 | 2000-2024 |
| COG | Congo | 2004 | 2011-2024 |
| COM | Comoros | 2004 | 2000-2004, 2006-2023 |
| DJI | Djibouti | 2004 | 2005-2014, 2016-2022 |
| ERI | Eritrea | 2004 | 2002-2024 |
| ETH | Ethiopia | 2004 | 2000-2024 |
| GAB | Gabon | 2005 | 2000-2008, 2013-2019, 2022 |
| GHA | Ghana | 2004 | 2000-2024 |
| GIN | Guinea | 2004 | 2002-2024 |
| GMB | Gambia | 2005 | 2004-2009, 2011-2024 |
| GNB | Guinea-Bissau | 2004 | 2005-2024 |
| GNQ | Equatorial Guinea | 2005 | 2007-2009, 2011-2019, 2023-2024 |
| KEN | Kenya | 2004 | 2001-2024 |
| LBR | Liberia | 2004 | 2009-2015, 2017-2019, 2021-2024 |
| MDG | Madagascar | 2004 | 2001-2024 |
| MLI | Mali | 2004 | 2005-2024 |
| MOZ | Mozambique | 2004 | 2000-2024 |
| MRT | Mauritania | 2004 | 2006, 2008-2021 |
| MWI | Malawi | 2005 | 2006-2024 |
| NAM | Namibia | 2005 | 2010-2015, 2018 |
| NER | Niger | 2004 | 2005-2022 |
| NGA | Nigeria | 2004 | 2001-2024 |
| RWA | Rwanda | 2006 | 2000-2007, 2009-2024 |
| SDN | Sudan | 2004 | 2001-2022 |
| SEN | Senegal | 2004 | 2004-2024 |
| SLE | Sierra-Leone | 2004 | 2005-2024 |
| SOM | Somalia | 2004 | 2004-2022 |
| SSD | South Sudan | 2004 | 2005-2024 |
| STP | Sao Tome and Principe | 2006 | 2010-2022 |
| SWZ | Eswatini | 2006 | 2010-2012, 2014-2016, 2022 |
| TCD | Chad | 2004 | 2006-2011, 2013-2024 |
| TGO | Togo | 2004 | 2002-2024 |
| TZA | Tanzania | 2005 | 2002-2024 |
| UGA | Uganda | 2004 | 2005-2024 |
| ZMB | Zambia | 2004 | 2002-2024 |
| ZWE | Zimbabwe | 2004 | 2007-2010, 2012-2024 |

##### 2.1.2 Subnational Data

One main change present in the MITN model is the added capability to ingest subnational distribution data. For several country-years (see Table 2), subnational (Admin 1) level ITN distribution data can be inferred from AMP campaign trackers. Whilst not necessarily accurate, they provide a reasonable prior estimate of subnational level distribution.

Disaggregation of national level distribution data to a subnational level is done according to the following steps:

1. For each subnational Admin 1 region $j$ in a country $i$ : If AMP subnational distribution data is available for a given country-Admin 1-year, then set the estimated subnational distribution $\hat{d}_{i,j}(t)$to be equal to the AMP tracker data. Otherwise, just disaggregate based the relative population fraction of each subnational region.
2. Calculate $\hat{d}_i(t) = \sum_j \hat{d}_{i,j}(t)$ to be the total national distributions based on the AMP tracker.
3. Check the following
   - (a) If the national estimate $\hat{d}_i(t)$ is less than the reported national distribution $\hat{d}(t)$, then assign the excess nets $d_i(t) - \hat{d}_i(t)$ amongst regions with no pre-existing subnational based on their relative population sizes.
   - (b) If $\hat{d}_i(t)$ is greater than the reported national distribution $\hat{d}(t)$, reduce all subnational distributions by a ratio such that they are equal.

Table 2: Data availability summary for subnational ITN distributions taken from AMP tracker.

| **ISO** | **Country** | **Admin 1** | **Year** |
|---|---|---|---|
| BDI | Burundi | Bubanza | 2022 |
| | | Bujumbura Mairie | 2022 |
| | | Bujumbura Rural | 2022 |
| | | Bururi | 2022 |
| | | Cankuzo | 2022 |
| | | Cibitoke | 2022 |
| | | Gitega | 2022 |
| | | Karuzi | 2022 |
| | | Kayanza | 2022 |
| | | Kirundo | 2022 |
| | | Makamba | 2022 |
| | | Muramvya | 2022 |
| | | Mwaro | 2022 |
| | | Rumonge | 2022 |
| | | Rutana | 2022 |
| | | Ruyigi | 2022 |
| BFA | Burkina Faso | Boucle du Mouhoun | 2022 |
| | | Centre | 2022 |
| | | Centre-Est | 2022 |
| | | Centre-Nord | 2022 |
| | | Centre-Ouest | 2022 |
| | | Centre-Sud | 2022 |
| | | Est | 2022 |
| | | Hauts-Bassins | 2022 |
| | | Nord | 2022 |
| | | Plateau Central | 2022 |
| CMR | Cameroon | Adamaoua | 2022 |
| | | Centre | 2022 |
| | | Est | 2022 |
| | | Extreme Nord | 2022 |
| | | Littoral | 2022 |
| | | Nord | 2022 |
| | | Ouest | 2022 |
| | | Sud | 2022 |
| | | Bas-Uele | 2021 |
| | | Haut-Uele | 2021 |
| | | Ituri | 2021 |



COD Democratic Republic of the Congo

| | | | |
|---|---|---|---|
| | | Kasai | 2022 |
| | | Kasai Central | 2021 |
| | | Kinshasa | 2021 |
| | | Kwango | 2021 |
| | | Kwilu | 2021 |
| | | Lualaba | 2021 |
| | | Maniema | 2021 |
| | | Nord-Kivu | 2021 |
| | | Sankuru | 2021 |
| | | Sud-Kivu | 2021 |
| | | Tshopo | 2021 |
| DJI | Djibouti | Djibouti | 2022 |
| ETH | Ethiopia | Afar | 2022 |
| | | Amhara | 2021 |
| | | Amhara | 2022 |
| | | Benishangul Gumz | 2022 |
| | | Dire Dawa | 2021 |
| | | Gambela | 2022 |
| | | Harari | 2021 |
| | | Oromia | 2021 |
| | | Oromia | 2022 |
| | | SNNP | 2022 |
| | | Sidama | 2022 |
| | | Somali | 2021 |
| | | Tigray | 2022 |
| GHA | Ghana | Ahafo | 2021 |
| | | Ashanti | 2021 |
| | | Bono | 2021 |
| | | Bono East | 2021 |
| | | Central | 2021 |
| | | Northern | 2021 |
| | | Northern East | 2021 |
| | | Oti | 2021 |
| | | Savannah | 2021 |
| | | Upper East | 2021 |
| | | Western North | 2021 |
| GIN | Guinea | Boke | 2022 |
| | | Conakry | 2022 |
| | | Faranah | 2022 |
| | | Kankan | 2022 |
| | | Kindia | 2022 |
| | | Labe | 2022 |
| | | Mamou | 2022 |
| | | Nzerekore | 2022 |
| GMB | Gambia | Central River | 2022 |
| | | Lower River | 2022 |
| | | North Bank | 2022 |
| | | Upper River | 2022 |
| | | West Coast | 2022 |
| KEN | Kenya | Baringo | 2021 |
| | | Bomet | 2021 |
| | | Bungoma | 2021 |
| | | Busia | 2021 |
| | | Homa Bay | 2021 |
| | | Kakamega | 2021 |
| | | Kericho | 2021 |
| | | Kilifi | 2021 |
| | | Kirinyaga | 2021 |
| | | Kisii | 2021 |
| | | Kisumu | 2021 |

| | | | |
|---|---|---|---|
| | | Kwale | 2021 |
| | | Lamu | 2021 |
| | | Marsabit | 2021 |
| | | Migori | 2021 |
| | | Mombasa | 2021 |
| | | Nandi | 2021 |
| | | Narok | 2021 |
| | | Nyamira | 2021 |
| | | Siaya | 2021 |
| | | Taita/taveta | 2021 |
| | | Tana River | 2021 |
| | | Trans Nzoia | 2021 |
| | | Turkana | 2021 |
| | | Uasin Gishu | 2021 |
| | | Vihiga | 2021 |
| | | West Pokot | 2021 |
| LBR | Liberia | Bomi | 2021 |
| | | Bong | 2021 |
| | | Gbapolu | 2021 |
| | | Grand Bassa | 2021 |
| | | Grand Cape Mount | 2021 |
| | | Grand Gedeh | 2021 |
| | | Grand Kru | 2021 |
| | | Lofa | 2021 |
| | | Margibi | 2021 |
| | | Maryland | 2021 |
| | | Nimba | 2021 |
| | | River Cess | 2021 |
| | | River Gee | 2021 |
| | | Sinoe | 2021 |
| MDG | Madagascar | Alaotra Mangoro | 2021 |
| | | Amoron I Mania | 2021 |
| | | Analamanga | 2021 |
| | | Analanjirofo | 2021 |
| | | Androy | 2021 |
| | | Anosy | 2021 |
| | | Atsimo Andrefana | 2021 |
| | | Atsimo Atsinanana | 2021 |
| | | Atsinanana | 2021 |
| | | Betsiboka | 2021 |
| | | Boeny | 2021 |
| | | Bongolava | 2021 |
| | | Diana | 2021 |
| | | Haute Matsiatra | 2021 |
| | | Ihorombe | 2021 |
| | | Melaky | 2021 |
| | | Menabe | 2021 |
| | | Sava | 2021 |
| | | Sofia | 2021 |
| | | Vakinankaratra | 2021 |
| | | Vatovavy Fitovinany | 2021 |
| MOZ | Mozambique | Cabo Delgado | 2022 |
| | | Niassa | 2022 |
| MWI | Malawi | Central Region | 2021 |
| | | Central Region | 2022 |
| | | Northern Region | 2021 |
| | | Northern Region | 2022 |
| | | Southern Region | 2021 |
| NER | Niger | Agadez | 2021 |
| | | Maradi | 2021 |

| | | | |
|---|---|---|---|
| | | Tahoua | 2021 |
| | | Zinder | 2021 |
| NGA | Nigeria | Delta | 2022 |
| | | Gombe | 2021 |
| | | Jigawa | 2021 |
| | | Kaduna | 2022 |
| | | Katsina | 2021 |
| | | Kebbi | 2021 |
| | | Nassarawa | 2021 |
| | | Nassarawa | 2022 |
| | | Niger | 2022 |
| | | Ogun | 2021 |
| | | Ondo | 2021 |
| | | Oyo | 2021 |
| | | Sokoto | 2021 |
| | | Taraba | 2022 |
| | | Yobe | 2022 |
| SEN | Senegal | Diourbel | 2022 |
| | | Fatick | 2022 |
| | | Kaffrine | 2022 |
| | | Kaolack | 2022 |
| | | Kedougou | 2022 |
| | | Kolda | 2022 |
| SEN | Senegal | Louga | 2022 |
| | | Matam | 2022 |
| | | Saint-Louis | 2022 |
| | | Sedhiou | 2022 |
| | | Tambacounda | 2022 |
| | | Ziguinchor | 2022 |
| SOM | Somalia | Bakool | 2022 |
| | | Banadir | 2022 |
| | | Bay | 2022 |
| | | Galgaduud | 2022 |
| | | Gedo | 2022 |
| | | Hiraan | 2022 |
| | | Juba Hoose | 2022 |
| | | Shabelle Hoose | 2022 |

#### 2.2 Household Survey Data

Household survey data pertaining to net ownership, access and use is used to calibrate the SNF model in order to identify plausible net attrition parameters. Two typs of household data is present: (1) Individual household survey with geolocation coordinates given up to a cluster level, and (2) population level aggregated values taken from government published summary reports. For the former case, household survey entries are provided at a monthly resolution and contains information on household size, number of nets owned and itn usage behaviour. For the latter, aggregates are attributed to the time midpoint of the reported start and end of the survey period. A summary of the household survey data availability by country is given in Tables 3 and 4.

Table 3: Summary of available household net ownership surveys used in analysis, and associated number of subnational Admin 1 regions.

| ISO | Country | Num. Household Surveys | Num. Reports | Num. Subnational Regions |
|---|---|---|---|---|
| AGO | Angola | 3 | 0 | 18 |
| BDI | Burundi | 3 | 0 | 19 |
| BEN | Benin | 4 | 0 | 12 |
| BFA | Burkina Faso | 5 | 0 | 13 |
| CAF | Ctr. Afr. Republic | 2 | 0 | 17 |
| CIV | Côte d'Ivoire | 4 | 0 | 14 |
| CMR | Cameroon | 5 | 0 | 10 |
| COD | Dem. Rep. Congo | 4 | 0 | 26 |
| COG | Congo | 3 | 0 | 12 |
| COM | Comoros | 1 | 0 | 3 |
| DJI | Djibouti | 0 | 1 | 5 |
| ETH | Ethiopia | 1 | 3 | 12 |
| GAB | Gabon | 2 | 0 | 9 |
| GHA | Ghana | 9 | 0 | 16 |
| GIN | Guinea | 5 | 0 | 8 |
| GMB | Gambia | 4 | 2 | 6 |
| GNB | Guinea-Bissau | 2 | 0 | 9 |
| GNQ | Equatorial Guinea | 0 | 1 | 7 |
| KEN | Kenya | 7 | 0 | 49 |
| LBR | Liberia | 6 | 0 | 15 |
| MDG | Madagascar | 7 | 0 | 22 |
| MLI | Mali | 7 | 0 | 9 |
| MOZ | Mozambique | 4 | 0 | 12 |
| MRT | Mauritania | 3 | 0 | 13 |
| MWI | Malawi | 8 | 1 | 4 |
| NAM | Namibia | 2 | 0 | 13 |
| NER | Niger | 3 | 0 | 8 |
| NGA | Nigeria | 9 | 0 | 37 |
| RWA | Rwanda | 7 | 0 | 6 |
| SEN | Senegal | 13 | 1 | 14 |
| SLE | Sierra Leone | 6 | 0 | 5 |
| SOM | Somalia | 1 | 0 | 18 |
| SSD | South Sudan | 0 | 3 | 10 |
| STP | Sao Tome and Principe | 3 | 0 | 2 |
| SWZ | Eswatini | 2 | 0 | 4 |
| TCD | Chad | 3 | 0 | 23 |
| TGO | Togo | 4 | 0 | 5 |
| TZA | Tanzania | 7 | 0 | 36 |
| UGA | Uganda | 6 | 0 | 5 |
| ZMB | Zambia | 4 | 3 | 10 |
| ZWE | Zimbabwe | 5 | 0 | 10 |

Table 4: Metadata summary of compiled ITN ownership dataset

| ISO | Country | Dataset ID | Data Type | Start Date | End Date | Sample Size |
|---|---|---|---|---|---|---|
| AGO | Angola | AO2006MIS | Household Survey | 2006-11 | 2007-3 | 2273 |
| AGO | Angola | AO2011MIS | Household Survey | 2010-2 | 2011-6 | 7111 |
| AGO | Angola | AO2015DHS | Household Survey | 2015-10 | 2016-3 | 14096 |
| BDI | Burundi | BU2010DHS | Household Survey | 2010-8 | 2011-1 | 7455 |
| BDI | Burundi | BU2012MIS | Household Survey | 2012-11 | 2013-1 | 4293 |
| BDI | Burundi | BU2016DHS | Household Survey | 2016-10 | 2017-3 | 14208 |
| BEN | Benin | BJ2006DHS | Household Survey | 2006-8 | 2006-11 | 17511 |
| BEN | Benin | BJ2012DHS | Household Survey | 2011-12 | 2012-4 | 15507 |
| BEN | Benin | BEN2014MICS | Household Survey | 2014-6 | 2014-9 | 14077 |
| BEN | Benin | BJ2017DHS | Household Survey | 2017-11 | 2018-2 | 12651 |
| BFA | Burkina Faso | BF2003DHS | Household Survey | 2003-6 | 2003-11 | 8501 |
| BFA | Burkina Faso | BF2010DHS | Household Survey | 2010-1 | 2010-12 | 12937 |
| BFA | Burkina Faso | BF2014MIS | Household Survey | 2014-9 | 2014-12 | 5688 |
| BFA | Burkina Faso | BF2017MIS | Household Survey | 2017-11 | 2018-3 | 5605 |
| BFA | Burkina Faso | BF2021DHS | Household Survey | 2021-7 | 2021-11 | 11481 |
| CAF | Ctr. Afr. Republic | CAF2010MICS | Household Survey | 2010-6 | 2010-12 | 11755 |
| CAF | Ctr. Afr. Republic | CAF2018MICS | Household Survey | 2018-11 | 2019-12 | 8133 |
| CIV | Côte d'Ivoire | CI2005AIS | Household Survey | 2005-8 | 2005-10 | 4342 |
| CIV | Côte d'Ivoire | CI2012DHS | Household Survey | 2011-12 | 2012-5 | 9462 |
| CIV | Côte d'Ivoire | CIV2016MICS | Household Survey | 2016-4 | 2016-7 | 11878 |
| CIV | Côte d'Ivoire | CI2021DHS | Household Survey | 2021-9 | 2021-12 | 14444 |
| CMR | Cameroon | CM2004DHS | Household Survey | 2004-2 | 2004-8 | 9429 |
| CMR | Cameroon | CM2011DHS | Household Survey | 2011-1 | 2011-8 | 6555 |
| CMR | Cameroon | CMR2014MICS | Household Survey | 2014-6 | 2014-10 | 10213 |
| CMR | Cameroon | CM2018DHS | Household Survey | 2018-6 | 2019-1 | 10189 |
| CMR | Cameroon | CM2022MIS | Household Survey | 2022-8 | 2022-11 | 5519 |
| COD | Dem Rep. Congo | CD2007DHS | Household Survey | 2007-2 | 2007-7 | 8676 |
| COD | Dem Rep. Congo | COD2010MICS | Household Survey | 2010-2 | 2010-4 | 11391 |
| COD | Dem Rep. Congo | CD2013DHS | Household Survey | 2013-8 | 2014-2 | 17796 |
| COD | Dem Rep. Congo | COD2017MICS | Household Survey | 2017-11 | 2018-7 | 20312 |
| COG | Congo | CG2005DHS | Household Survey | 2005-7 | 2005-11 | 5878 |
| COG | Congo | CG2011DHS | Household Survey | 2011-1 | 2012-2 | 11581 |
| COG | Congo | COG2014MICS | Household Survey | 2014-11 | 2015-2 | 12811 |
| COM | Comoros | KM2012DHS | Household Survey | 2012-8 | 2012-12 | 3670 |
| DJI | Djibouti | REP_DJI2008MIS | Report Summary | 2009-1 | 2009-1 | 1 |
| ETH | Ethiopia | ET2005DHS | Household Survey | 2005-4 | 2005-8 | 12280 |
| ETH | Ethiopia | REP_ETH2007MIS | Report Summary | 2007-11 | 2007-11 | 1 |
| ETH | Ethiopia | REP_ETH2011MIS | Report Summary | 2011-11 | 2011-11 | 1 |
| ETH | Ethiopia | REP_ETH2015MIS | Report Summary | 2015-11 | 2015-11 | 1 |
| GAB | Gabon | GA2012DHS | Household Survey | 2012-1 | 2012-5 | 8668 |
| GAB | Gabon | GA2019DHS | Household Survey | 2019-11 | 2021-10 | 10291 |
| GHA | Ghana | GH2003DHS | Household Survey | 2003-7 | 2003-10 | 5470 |
| GHA | Ghana | GH2008DHS | Household Survey | 2008-9 | 2008-11 | 10458 |
| GHA | Ghana | 2010MICS | Household Survey | 2010-12 | 2011-1 | 1409 |
| GHA | Ghana | GHA2011MICS | Household Survey | 2011-9 | 2011-12 | 11925 |
| GHA | Ghana | GH2014DHS | Household Survey | 2014-9 | 2014-12 | 10365 |
| GHA | Ghana | GH2016MIS | Household Survey | 2016-10 | 2016-11 | 5129 |
| GHA | Ghana | GHA2017MICS | Household Survey | 2017-10 | 2018-1 | 12884 |
| GHA | Ghana | GH2019MIS | Household Survey | 2019-9 | 2019-11 | 5240 |
| GHA | Ghana | GH2022DHS | Household Survey | 2022-10 | 2023-1 | 16184 |
| GIN | Guinea | GN2005DHS | Household Survey | 2005-2 | 2005-6 | 5745 |
| GIN | Guinea | GN2012DHS | Household Survey | 2012-6 | 2012-10 | 6340 |
| GIN | Guinea | GIN2016MICS | Household Survey | 2016-8 | 2016-11 | 8080 |
| GIN | Guinea | GN2018DHS | Household Survey | 2018-3 | 2018-6 | 6580 |
| GIN | Guinea | GN2021MIS | Household Survey | 2021-7 | 2021-9 | 3600 |
| GMB | Gambia | GMB2010MICS | Household Survey | 2010-4 | 2010-8 | 7791 |

| | | | | | | |
|---|---|---|---|---|---|---|
| GMB | Gambia | GM2013DHS | Household Survey | 2013-2 | 2013-4 | 6216 |
| GMB | Gambia | REP_GMB2014MIS | Report Summary | 2014-12 | 2014-12 | 1 |
| GMB | Gambia | REP_GMB2017MIS | Report Summary | 2017-12 | 2017-12 | 1 |
| GMB | Gambia | GMB2018MICS | Household Survey | 2018-1 | 2018-4 | 7405 |
| GMB | Gambia | GM2019DHS | Household Survey | 2019-11 | 2020-3 | 5940 |
| GNB | Guinea-Bissau | GNB2014MICS | Household Survey | 2014-3 | 2014-7 | 6601 |
| GNB | Guinea-Bissau | GNB2018MICS | Household Survey | 2018-11 | 2019-3 | 7377 |
| GNQ | Equatorial Guinea | REP_GNQ2008DHS | Report Summary | 2011-9 | 2011-9 | 1 |
| KEN | Kenya | KE2003DHS | Household Survey | 2003-4 | 2003-9 | 7457 |
| KEN | Kenya | KE2008DHS | Household Survey | 2008-10 | 2009-3 | 8001 |
| KEN | Kenya | 2013MICS | Household Survey | 2013-11 | 2014-2 | 3743 |
| KEN | Kenya | KE2014DHS | Household Survey | 2014-4 | 2014-10 | 32164 |
| KEN | Kenya | KE2015MIS | Household Survey | 2015-7 | 2015-8 | 5462 |
| KEN | Kenya | KE2020MIS | Household Survey | 2020-11 | 2020-12 | 6842 |
| KEN | Kenya | KE2022DHS | Household Survey | 2022-2 | 2022-7 | 34693 |
| LBR | Liberia | LB2009MIS | Household Survey | 2008-12 | 2009-3 | 3637 |
| LBR | Liberia | LB2011MIS | Household Survey | 2011-9 | 2011-12 | 3733 |
| LBR | Liberia | LB2013DHS | Household Survey | 2013-3 | 2013-7 | 8500 |
| LBR | Liberia | LB2016MIS | Household Survey | 2016-9 | 2016-11 | 3743 |
| LBR | Liberia | LB2019DHS | Household Survey | 2019-10 | 2020-2 | 7999 |
| LBR | Liberia | LB2022MIS | Household Survey | 2022-10 | 2022-12 | 3879 |
| MDG | Madagascar | MD2008DHS | Household Survey | 2008-11 | 2009-7 | 15954 |
| MDG | Madagascar | MD2011MIS | Household Survey | 2011-3 | 2011-5 | 7157 |
| MDG | Madagascar | 2012MICS | Household Survey | 2012-5 | 2012-7 | 2967 |
| MDG | Madagascar | MD2013MIS | Household Survey | 2013-4 | 2013-6 | 7494 |
| MDG | Madagascar | MD2016MIS | Household Survey | 2016-4 | 2016-7 | 9962 |
| MDG | Madagascar | MDG2018MICS | Household Survey | 2018-8 | 2018-12 | 17870 |
| MDG | Madagascar | MD2021DHS | Household Survey | 2021-3 | 2021-7 | 18372 |
| MLI | Mali | ML2006DHS | Household Survey | 2006-4 | 2006-12 | 11399 |
| MLI | Mali | MLI2009MICS | Household Survey | 2009-12 | 2010-8 | 13848 |
| MLI | Mali | ML2012DHS | Household Survey | 2012-11 | 2013-2 | 8810 |
| MLI | Mali | MLI2015MICS | Household Survey | 2015-7 | 2015-10 | 11830 |
| MLI | Mali | ML2015MIS | Household Survey | 2015-9 | 2015-11 | 3868 |
| MLI | Mali | ML2018DHS | Household Survey | 2018-8 | 2018-11 | 8336 |
| MLI | Mali | ML2021MIS | Household Survey | 2021-9 | 2021-11 | 5040 |
| MOZ | Mozambique | MZ2011DHS | Household Survey | 2011-5 | 2011-12 | 13080 |
| MOZ | Mozambique | MZ2015AIS | Household Survey | 2015-5 | 2015-9 | 6253 |
| MOZ | Mozambique | MZ2018MIS | Household Survey | 2018-3 | 2018-6 | 5335 |
| MOZ | Mozambique | MZ2022DHS | Household Survey | 2022-7 | 2023-2 | 12948 |
| MRT | Mauritania | MRT2011MICS | Household Survey | 2011-6 | 2011-11 | 10112 |
| MRT | Mauritania | MRT2015MICS | Household Survey | 2015-7 | 2015-11 | 11765 |
| MRT | Mauritania | MR2020DHS | Household Survey | 2019-11 | 2021-3 | 10742 |
| MWI | Malawi | MW2004DHS | Household Survey | 2004-1 | 2005-2 | 12060 |
| MWI | Malawi | REP_MWI2010MIS | Report Summary | 2010-4 | 2010-4 | 1 |
| MWI | Malawi | MW2010DHS | Household Survey | 2010-6 | 2010-9 | 22145 |
| MWI | Malawi | MW2012MIS | Household Survey | 2012-3 | 2012-4 | 3133 |
| MWI | Malawi | MWI2013MICS | Household Survey | 2013-11 | 2014-4 | 26712 |
| MWI | Malawi | MW2014MIS | Household Survey | 2014-5 | 2014-6 | 3015 |
| MWI | Malawi | MW2015DHS | Household Survey | 2015-10 | 2016-2 | 23102 |
| MWI | Malawi | MW2017MIS | Household Survey | 2017-4 | 2017-6 | 3152 |
| MWI | Malawi | MWI2019MICS | Household Survey | 2019-12 | 2020-8 | 25418 |
| NAM | Namibia | NM2006DHS | Household Survey | 2006-11 | 2007-3 | 8346 |
| NAM | Namibia | NM2013DHS | Household Survey | 2013-4 | 2013-9 | 9066 |
| NER | Niger | NI2006DHS | Household Survey | 2006-1 | 2006-6 | 7659 |
| NER | Niger | NI2012DHS | Household Survey | 2012-1 | 2012-6 | 10747 |
| NER | Niger | NI2021MIS | Household Survey | 2021-8 | 2021-10 | 4314 |
| NGA | Nigeria | NG2003DHS | Household Survey | 2003-3 | 2003-8 | 6267 |
| NGA | Nigeria | NG2008DHS | Household Survey | 2008-6 | 2008-11 | 30699 |

| | | | | | | |
|---|---|---|---|---|---|---|
| NGA | Nigeria | NG2010MIS | Household Survey | 2010-10 | 2010-12 | 5092 |
| NGA | Nigeria | NGA2011MICS | Household Survey | 2011-2 | 2011-4 | 29074 |
| NGA | Nigeria | NG2013DHS | Household Survey | 2013-2 | 2013-7 | 33924 |
| NGA | Nigeria | NG2015MIS | Household Survey | 2015-10 | 2015-11 | 6869 |
| NGA | Nigeria | NGA2016MICS | Household Survey | 2016-9 | 2017-1 | 33901 |
| NGA | Nigeria | NG2018DHS | Household Survey | 2018-8 | 2018-12 | 35894 |
| NGA | Nigeria | NG2021MIS | Household Survey | 2021-10 | 2021-12 | 12658 |
| RWA | Rwanda | RW2005DHS | Household Survey | 2005-2 | 2005-7 | 8973 |
| RWA | Rwanda | RW2008DHS | Household Survey | 2007-12 | 2008-4 | 6388 |
| RWA | Rwanda | RW2010DHS | Household Survey | 2010-9 | 2011-3 | 11206 |
| RWA | Rwanda | RW2013MIS | Household Survey | 2013-2 | 2013-4 | 4766 |
| RWA | Rwanda | RW2015DHS | Household Survey | 2014-11 | 2015-4 | 11493 |
| RWA | Rwanda | RW2017MIS | Household Survey | 2017-10 | 2017-12 | 5025 |
| RWA | Rwanda | RW2019DHS | Household Survey | 2019-11 | 2020-7 | 11593 |
| SEN | Senegal | SN2005DHS | Household Survey | 2005-1 | 2005-6 | 6705 |
| SEN | Senegal | SN2006MIS | Household Survey | 2006-6 | 2006-12 | 3061 |
| SEN | Senegal | SN2008MIS | Household Survey | 2008-1 | 2009-2 | 9857 |
| SEN | Senegal | SN2010DHS | Household Survey | 2010-10 | 2011-4 | 7106 |
| SEN | Senegal | SN2012DHS | Household Survey | 2012-9 | 2013-6 | 3805 |
| SEN | Senegal | REP_SEN2012DHS | Report Summary | 2013-2 | 2013-2 | 1 |
| SEN | Senegal | SN2014DHS | Household Survey | 2014-1 | 2014-10 | 3680 |
| SEN | Senegal | 2015MICS | Household Survey | 2015-10 | 2016-1 | 4948 |
| SEN | Senegal | SN2015DHS | Household Survey | 2015-2 | 2015-11 | 3944 |
| SEN | Senegal | SN2016DHS | Household Survey | 2016-3 | 2016-11 | 3786 |
| SEN | Senegal | SN2017DHS | Household Survey | 2017-4 | 2017-12 | 7549 |
| SEN | Senegal | SN2018DHS | Household Survey | 2018-5 | 2018-12 | 4085 |
| SEN | Senegal | SN2019DHS | Household Survey | 2019-4 | 2019-12 | 4214 |
| SEN | Senegal | SN2020MIS | Household Survey | 2020-12 | 2021-1 | 4687 |
| SLE | Sierra Leone | SL2008DHS | Household Survey | 2008-4 | 2008-6 | 6162 |
| SLE | Sierra Leone | SLE2010MICS | Household Survey | 2010-10 | 2010-12 | 11393 |
| SLE | Sierra Leone | SL2013DHS | Household Survey | 2013-6 | 2013-11 | 11238 |
| SLE | Sierra Leone | SL2016MIS | Household Survey | 2016-6 | 2016-8 | 5806 |
| SLE | Sierra Leone | SLE2017MICS | Household Survey | 2017-5 | 2017-8 | 15309 |
| SLE | Sierra Leone | SL2019DHS | Household Survey | 2019-5 | 2019-8 | 11746 |
| SOM | Somalia | 2011MICS | Household Survey | 2011-4 | 2011-12 | 16376 |
| SSD | South Sudan | REP_SSD2009MIS | Report Summary | 2009-12 | 2009-12 | 1 |
| SSD | South Sudan | REP_SSD2013MIS | Report Summary | 2013-10 | 2013-10 | 1 |
| SSD | South Sudan | REP_SSD2017MIS | Report Summary | 2017-12 | 2017-12 | 1 |
| STP | Sao Tome and Principe | ST2008DHS | Household Survey | 2008-1 | 2009-1 | 3536 |
| STP | Sao Tome and Principe | STP2014MICS | Household Survey | 2014-4 | 2014-6 | 3492 |
| STP | Sao Tome and Principe | STP2019MICS | Household Survey | 2019-7 | 2019-10 | 3426 |
| SWZ | Eswatini | SZ2006DHS | Household Survey | 2006-7 | 2007-2 | 4374 |
| SWZ | Eswatini | SWZ2010MICS | Household Survey | 2010-8 | 2010-11 | 4834 |
| TCD | Chad | TCD2010MICS | Household Survey | 2010-1 | 2010-5 | 16385 |
| TCD | Chad | TD2014DHS | Household Survey | 2014-10 | 2015-4 | 9932 |
| TCD | Chad | TCD2019MICS | Household Survey | 2019-4 | 2019-12 | 18958 |
| TGO | Togo | TGO2010MICS | Household Survey | 2010-9 | 2010-11 | 6038 |
| TGO | Togo | TG2013DHS | Household Survey | 2013-1 | 2014-4 | 8343 |
| TGO | Togo | TGO2017MICS | Household Survey | 2017-7 | 2017-10 | 7916 |
| TGO | Togo | TG2017MIS | Household Survey | 2017-9 | 2017-11 | 4478 |
| TZA | Tanzania | TZ2004DHS | Household Survey | 2004-10 | 2005-2 | 9686 |
| TZA | Tanzania | TZ2007AIS | Household Survey | 2007-10 | 2008-2 | 7707 |
| TZA | Tanzania | TZ2010DHS | Household Survey | 2009-12 | 2010-5 | 8839 |
| TZA | Tanzania | TZ2012AIS | Household Survey | 2011-12 | 2012-5 | 8565 |
| TZA | Tanzania | TZ2015DHS | Household Survey | 2015-1 | 2016-2 | 10989 |
| TZA | Tanzania | TZ2017MIS | Household Survey | 2017-10 | 2017-12 | 7873 |
| TZA | Tanzania | TZ2022DHS | Household Survey | 2022-2 | 2022-7 | 14051 |
| UGA | Uganda | UG2006DHS | Household Survey | 2006-5 | 2006-10 | 8065 |

| | | | | | | |
|---|---|---|---|---|---|---|
| UGA | Uganda | UG2009MIS | Household Survey | 2009-11 | 2010-2 | 3961 |
| UGA | Uganda | UG2011DHS | Household Survey | 2011-6 | 2011-12 | 7578 |
| UGA | Uganda | UG2014MIS | Household Survey | 2014-12 | 2015-2 | 4690 |
| UGA | Uganda | UG2016DHS | Household Survey | 2016-6 | 2016-12 | 17092 |
| UGA | Uganda | UG2018MIS | Household Survey | 2018-12 | 2019-1 | 7308 |
| ZMB | Zambia | ZM2002DHS | Household Survey | 2001-11 | 2002-6 | 7125 |
| ZMB | Zambia | ZM2007DHS | Household Survey | 2007-4 | 2007-10 | 6577 |
| ZMB | Zambia | REP_ZMB2008MIS | Report Summary | 2008-4 | 2008-4 | 1 |
| ZMB | Zambia | REP_ZMB2012MIS | Report Summary | 2012-4 | 2012-4 | 1 |
| ZMB | Zambia | ZM2013DHS | Household Survey | 2013-8 | 2014-4 | 13854 |
| ZMB | Zambia | REP_ZMB2015MIS | Report Summary | 2015-5 | 2018-4 | 2 |
| ZMB | Zambia | ZM2018DHS | Household Survey | 2018-7 | 2019-1 | 11124 |
| ZWE | Zimbabwe | ZW2005DHS | Household Survey | 2005-8 | 2006-4 | 8125 |
| ZWE | Zimbabwe | ZW2010DHS | Household Survey | 2010-9 | 2011-3 | 8552 |
| ZWE | Zimbabwe | ZWE2014MICS | Household Survey | 2014-2 | 2014-4 | 15630 |
| ZWE | Zimbabwe | ZW2015DHS | Household Survey | 2015-7 | 2015-12 | 9151 |
| ZWE | Zimbabwe | ZWE2019MICS | Household Survey | 2018-12 | 2019-4 | 11091 |

### 3 Stock-and-Flow (SNF)

#### 3.1 National SNF

We first define the following variables and associated notation:

- $m_n$ : Reported number of LLINs delivered at the start of year $n$
- $d_n^{LLIN}$ : Reported number of LLINs distributed at the start of year $n$
- $\omega_n^+$: Number of LLINs in the national stock at the start of year $n$
- $\omega_n^-$: Number of LLINs in the national stock at the end of year $n$
- $\hat{\delta}_y^{LLIN}$: Number of LLINs distributed excluding additional redistribution of current excess net stock
- $\delta_y^{LLIN}$: Number of actual LLINs distributed during the entirety of year $n$
- $\phi$: Unique redistribution constant for each country.
- $P(t)$: the country population at month $t$
- $\gamma(t)$: the observed true nets-per-capita (NPC) estimated from household surveysmonth $t$
- $\Gamma(t) = \gamma(t)P(t)$: the estimated observed net crop in a given country.

Given a set of national delivery data $m_n^{LLIN}$, reported distribution data $m_n$ and observed net crop $\Gamma_t$ (where $n$ is years, $t$ is months), the general stock-and-flow SNF model for country possessing both LLINs and cITNs is given by the following set of difference equations as stated in the Methods section:

**Evolution Equations**

$$
\begin{aligned}
\omega_n^+ &= \omega_{n-1}^- + m_n \\
\hat{\delta}_n^{LLIN} &= \min(\omega_n^+, d_n^{LLIN}) \\
\delta_n^{LLIN} &= \hat{\delta}_n^{LLIN} + \phi(\omega_n^+ - \hat{\delta}_y^{LLIN}) \\
\omega_n^- &= \omega_n^+ - \delta_n^{LLIN}
\end{aligned}
$$

subject to the initial condition,

$$
\omega_0^+ = m_y,
$$

**LLIN-cITN Conversion**
To account for the lack of information regarding the number of distributed cITNs, an additional model parameter is introduced to convert between the two:

$$
\delta_n^{cITN} = \frac{\delta_n^{LLIN}}{\alpha^{LLIN}}.
$$

**Temporal Disaggregation**
SNF models for ITNs also face the problem of having disparate sampling frequencies in their input datasets. Specifically, delivery and distribution data is relatively consistent and reported at annual frequencies. In constrast, household survey against which net crop estimates need to be calibrated against often presented at a monthly but episodic frequency corresponding to the large length of time required to complete each round of nationally representative surveys. As described by [2], these disparate sampling frequencies can be reconciled using a set of learned proportion parameters,

$$\delta_{n,m} = \rho_{n,m}\delta_n,$$

where $\rho_{n,m}$ is the proportion of nets in the $n^{\text{th}}$ year that are distributed in the $m^{\text{th}}$ month of that year, and is subject to the constraint,

$$\sum_{m=1}^{12} p_{n,m} = 1, \forall n.$$

**Net Attrition**
Net attrition refers to the removal of nets from the observed net crop of a population that cause of which is non-specific and can be attributed to a variety of factors such as natural wear and tear, disposal, random losses or improper usage. The Bhatt model [3] originally proposed the usage of a two parameter compact exponential sigmoidal function to model the natural attrition of net crop over time. This approach was later simplified in the Bhatt-BV model by arbitrarily setting the shape parameter $\kappa = 20$, resulting in a lower number of parameters. Later findings from [2] suggest that a two parameter sigmoidal function is more appropriate for SNF models due to its flexibility, and does not impose a significant cost on parameter identifiability. Therefore, our analyses use the same attrition function proposed by Bhatt et al.,

$$f(t) = \begin{cases} e^{\kappa\left(1-\frac{1}{1-\left(\frac{t}{12\tau}\right)^2}\right)}, & t \in (0, 12\tau), \\ 0, \quad t \notin (0, 12\tau), \end{cases}$$

where $t$ is given in months. The model estimated total net crop $\hat{\Gamma}(t)$ at any given time $t$ is thus given as the convolution between the distribution time series, and the attrition function:

$$\hat{\Gamma}^{cITN}(t) = \langle \delta^{cITN} \star f^{cITN} \rangle(t) = \int_0^t f^{cITN}(T)\, \delta^{cITN}(t-T)\, dT,$$
$$\hat{\Gamma}^{LLIN}(t) = \langle \delta^{LLIN} \star f^{LLIN} \rangle(t) = \int_0^t f^{LLIN}(T)\, \delta^{LLIN}(t-T)\, dT.$$
$$\hat{\Gamma}(t) = \hat{\Gamma}^{cITN}(t) + \hat{\Gamma}^{LLIN}(t)$$

**Regression Equations**
Given the above governing equations, fitting the SNF model requires the calibration of model net-crop estimates against observed net crop estimates from household surveys given a set of input delivery and distribution time series. Thus, the calibration problem reduces to fitting,

$$\Gamma(t) \sim N(\hat{\Gamma}(t), \sigma^2(t)),$$

where $\sigma(t)$ is an inflated uncertainty derived from the sample standard error that accounts for volatility between months. Methods of calculation are found in [2]. To account for the large number of parameters in the model, calibration also requires the simultaneous minimisation of the loss function:

$$\mathcal{L}(\theta, \rho_t) = -2\mathbb{E}[\log(p(\hat{\Gamma}_t|\theta, \rho_t))] + \lambda \left\| \frac{\partial^2 \rho_t}{\partial t^2} \right\|^2,$$

such that temporal disaggregation parameters $\rho_t$ are not overfit.

Fitting of the entire model is done using an iterative expectation-maximisation (EM) algorithm as recommended by [2] and is demonstrated to be relatively robust convergence. The national SNF model can easily be extended to account for multiple LLIN types $i \in [PBO, G2, ROYAL, \dots]$ by altering the governing equations to the following:

$$\delta_n^{LLIN} = \sum_i \delta_n^{(i)}$$
$$\hat{\Gamma}^{(i)}(t) = \langle \delta^{(i)} \star (f^{(i)}|\tau_i, \kappa_i) \rangle(T) = \int_0^t f^{(i)}(T|\tau_i, \kappa_i)\, \delta^{(i)}(t-T)\, dT$$
$$\hat{\Gamma}^{LLIN}(t) = \sum_i \hat{\Gamma}^{(i)}(t)$$

##### 3.2 Subnational SNF

Similar to the national SNF model, a simplified subnational variant can also be fit given a set of subnational distribution time series and household observed estimates of net crop. In this case, the main simplification is the removal of the gating system based on the national deliveries and the assumption that estimated subnational distributions are sufficiently accurate. The national SNF can thus be adapted to construct estimates of net crop $\hat{\Gamma}_j^{(i)}(t)$ for subnational region $j$, with it's corresponding unique attrition parameter estimates $\tau_{subnat,j}, \kappa_{subnat,j}$. Furthermore, it is assumed that monthly distribution parameters $\rho_t$ for the subnational models are inherited from the national model.

Due to variations and potential inaccuracies in the subnational distribution data, in almost all cases national estimates of net crop $\hat{\Gamma}_{nat}(t)$ will differ from those arriving from the sum of subnational SNF estimates $\hat{\Gamma}_{subnat}(t) = \sum_i \sum_j \hat{\Gamma}_j^{(i)}(t)$. Here, we assume that national estimates are more reliable owing to their inherently larger sample size, and that distribution input time series are originally reported at a national level, the direct outputs from the subnational SNF are subsequently adjusted using three separate regressions:

**Conservation of Net Distributions** National total net distributions should be equal to the sum of subnational net distributions at all times,

$$\delta_{nat}(t) \sim N(\sum_j \omega_j \delta_{subnat,j}(t), (\epsilon \delta_{nat}(t))^2),$$

where $\omega_j$ are distribution weight adjustment parameters to be regressed, and $\epsilon$ is a parameter controlling the degree of allowable error.

**Conservation of Net Crop** National estimated net crop should closely match the sum of subnational net crop,

$$\hat{\Gamma}_{nat}(t) \sim N(\sum_j \nu_j \hat{\Gamma}_j(t), \sigma^2(t)),$$

where $\nu_j$ are crop weight adjustment parameters.

**Regression against Subnational Survey Observations**

$$\Gamma_j(t) \sim N(\hat{\Gamma}_j(t)|(\omega_j, \nu_j), \sigma^2_{subnat}(t)),$$

where $\sigma^2_{subnat}(t)$ is the related inflated uncertainty calcualted from the subnational subset of household survey observations.

### 4 Spatiotemporal Disaggregation

#### 4.1 Arithmetic (Gap) Normalisation

Disaggregation of national/subnational aggregates over a region into a set of values defined over a domain of smalle resolution is done by regressing the observed spatiotemporal values against a model consisting of the sum explanatory spatiotemporal covariates, combined with a Gaussian random field to account for model residuals that may be spatially correlated. As proposed by Bertozzi-Villa et al., such a regression can be easily implemented using the R-INLA package.

Prior to performing spatial disaggregation via INLA, local survey measures of ITN coverage (npc, access, use) cannot be directly used as response variables as they contain country specific factors that are not captured by covariates (e.g. net distribution behaviours, etc.). In order to construct a global spatio-temporal model that fits ITN coverage across the entire region, response variables must first be normalised such that any two points in the spatial domain may be directly compared against each other. The transformed response variable should ideally capture the relationship between local deviations in ITN coverage and topographical variation in covariates.

Previously, the Bhatt-BV model used several gap metrics in an attempt to define a normalised response variable. Instead of modelling NPC, access and use rates directly, response variables are defined as arithmetic deviations (the gap) from an aggregated baseline value. Specifically, the Bhatt-BV model uses national stock and flow estimates of NPC and access as the baseline value. The response variable for INLA is then defined as deviations from the national mean:

**NPC Gap**

$$\delta_\gamma(x) = \gamma(x) - \bar{\gamma}$$

**Access Gap**

$$\delta_\lambda(x) = \lambda(x) - \bar{\lambda}$$

where $\bar{\gamma}$ and $\bar{\lambda}$ are the mean stock and flow estimates of national NPC and access.

**Use Gap** Similarly, the response variable to represent use rate $\nu$ is also a gap metric, termed the use gap, but is normalised against the local access level instead because the stock and flow model does not account for conversions from access to use,

$$\delta_\nu(x) = \nu(x) - \lambda(x)$$

The domain for all three gap metrics is restricted to the interval $[-1, 1]$. For regression, gap metrics are first transformed via a logit link function to expand the domain to $[-\infty, \infty]$, followed by an inverse hyperbolic sine transform to minimise the effect of extreme values inherent in the range of the logistic function. For each case, the inverse hyperbolic sine transform is done with respect to a constant scaling parameter $\rho$ that is chosen such that the resultant log likelihood function arising from a Gaussian regression is as concentrated as possible,

$$\text{IHS}(x, \theta) = \frac{\text{arcsinh}\,(x/\theta)}{\theta}$$

where $\rho$ is chosen to maximise the following expression,

$$\mathcal{L}(\{x_i\}_{i=1}^n, \theta) = -n \log\left(\sum_{i=1}^n (x_i - \mathbb{E}[x]_i)^2\right) - \sum_{i=1}^n \log(1 + \theta^2 x_i^2)$$

The Bhatt-BV formulation of deviation response variables using an arithmetic gap is convenient for several reasons. Firstly, at allows for simple calculation of the final output rasters of $\gamma(x,t)$, $\lambda(x,t)$ and $\nu(x,t)$ using an element wise addition of matrices. Secondly, the simplicity of normalisation should limit the maximum size of the uncertainty. However, this transformation has three main inadequacies:

1. Arithmetic gaps do not guarantee that final interpolated estimates of local NPC, access and use are constrained to the appropriate domain. (i.e. $\gamma(x) \in [0, \infty]$, $\lambda(x), \nu(x) \in [0, 1]$)
2. Does not account for correlation between gap metrics and regional (e.g. national) stock and flow estimates.
3. Is not a proper normalisation and is not non-dimensional.

#### 4.2 Ratio Normalisation

A simple method to address the flaws associated with the arithmetic gap normalisation would be normalise using ratios with the baseline instead. Such an approach guarantees that normalised quantifies strictly adhere to the domain constraints of NPC, access and use once the unnormalised, which is important when attempting to reconstruct final maps of ITN coverage. Furthermore, taking the ratio against baseline values ensures that normalised quantities are properly non-dimensional. To this end, the MITN model instead uses three different normalised local deviation metrics:

**NPC deviation** ($\hat{\gamma}$)

$$\hat{\gamma} = \log\left(\frac{\gamma(x)}{\bar{\gamma}}\right)$$

**Deployment Rate** ($\hat{\psi}$)

$$\psi(x) = \frac{\lambda(x)}{2\gamma(x)}, \quad \bar{\psi} = \frac{\bar{\lambda}}{2\bar{\gamma}}$$

$$\hat{\psi} = \frac{\sinh^{-1}[g \circ h(\psi(x), \bar{\psi}))]}{\theta_\psi}$$

**Use-Access Deviation**($\hat{\xi}$)

$$\hat{\xi} = \frac{\theta_\xi \cdot \sinh^{-1}[g \circ h(\xi(x), \lambda(x)))]}{\theta_\xi}$$

For notation, bars represent SNF subnational aggregate estimates, and $x$ are spatial locations.. Functions $g, h$ are transformations used to ensure the correct constraints for domains are adhered to (i.e. $\gamma(x) \in [0, \infty)$, and $\lambda(x), \xi(x) \in [0, 1]$). These are defined as follows,

$$h(x, \mu) = \begin{cases} x^2, & \mu = 0 \\ -\left(\frac{|x-\mu|}{\mu}\right)^2, & 0 \le x < \mu < 1 \\ \left(\frac{|x-\mu|}{1-\mu}\right)^2, & 0 < \mu < x \le 1 \\ -(1-x)^2, & \mu = 1 \end{cases}$$

$$g(x) = \log\left(\frac{x/2 + 0.5}{1.5 - x/2}\right),$$

where $h$ is an asymmetric function that maps the deviation of an observation $x \in [0,1]$ from a reference point $\mu \in [0,1]$, and $g$ is a logit function with a transformation applied such that no special bias is given to observed values of $x = 0.5$.

To apply the above normalisation to household surveys, observations are first aggregated according to geolocated clusters to yield a set of geolocated observations of ITN coverage in time $\gamma(x,t), \lambda(x,t)$ and $\xi(x,t)$, where $x$ is a position corresponding to longitude/latitude coordinates. The above normalisation can then be applied to each cluster observation to yield normalised deviation values at each space-time coordinate $\hat{\gamma}(x,t), \hat{\psi}(x,t)$ and $\hat{\xi}(x,t)$. Finally, the following spatiotemporal model with random effects can be fit using R-INLA,

$$\hat{\gamma}(x,t) \sim \vec{\beta}_{\gamma,\,static} \cdot \vec{Y}_{static}(x) + \vec{\beta}_{\gamma,\,annual} \cdot \vec{Y}_{annual}(x) + F_{\gamma}(x,t),$$
$$\hat{\psi}(x,t) \sim \vec{\beta}_{\psi,\,static} \cdot \vec{Y}_{static}(x) + \vec{\beta}_{\psi,\,annual} \cdot \vec{Y}_{annual}(x) + F_{\psi}(x,t),$$
$$\hat{\xi}(x,t) \sim \vec{\beta}_{\xi,\,static} \cdot \vec{Y}_{static}(x) + \vec{Y}_{\xi,\,annual} \cdot \vec{Y}_{annual}(x) + \vec{Y}_{\xi,\,monthly} \cdot \vec{Y}_{monthly}(x) + F_{\xi}(x,t),$$

where $\vec{Y}$ are spatial/spatiotemporal covariates and $F$ is a spatiotemporal random field. The latter has a spatial component represented as a Gaussian process with Matérn covariance, and temporal component given by a first order autoregressive process. Whilst the above approach is performant and does yield correctly domain constrained values, the random field effect in the NPC model can resulted in greatly inflated values of local estimated NPC $\gamma(x,t)$ as it's domain is only constrained below. This effect can be partially mitigated by applying the random field for NPC to additive to the unnormalised observations instead,

$$\hat{\gamma}(x,t) \sim \vec{\beta}_{\gamma,\,static} \cdot \vec{Y}_{static}(x) + \vec{\beta}_{\gamma,\,annual} \cdot \vec{Y}_{annual}(x)$$
$$\gamma(x) - \bar{\gamma}e^{\hat{\gamma}} \sim F_{\gamma}(x,t).$$

### 5 Model Validation

The two-stage approach separating the SNF and spatiotemporal components employed by the Bhatt-BV and MITN model inherently produces two different estimates of NPC and access. These estimates correspond to (1) the SNF subnational aggregates, and (2) raster aggregates calculated by integrating over the spatially disaggregated ITN coverage rasters. The disparity between these estimates are the result of additional detail inferred by the spatial disaggregation model in regions where household surveys were not recorded.

Whilst sampling of households in surveys are designed to be demographically representative of each country [4], it is plausible that sampling is not necessarily geographically representative as some regions may be omitted in surveys due to logistical and cost constraints. Therefore, we argue that the detail and subsequent aggregate measures derived from spatiotemporal estimates provides a richer description of ITN coverage and should be used in preference to SNF estimates where possible unless continuity between previous national estimates of ITN coverage is required for comparison. We also note that the raking spatiotemporal aggregates against SNF estimates is not trivial as this requires applying nonlinear conservation constraints on the regressed SPDE spatial model.

To assess the model fit, both the SNF and spatiotemporal outputs of the MITN model are validated in isolation and together. Validation of the SNF model fit is quantified using calculated RMSE against the national survey aggregate estimate for each month. The fitted MITN model achieved an RMSE of 0.071 for NPC values, and 0.066 for access (see Figure 2). Validation of the spatiotemporal is done by comparing cluster level estimates NPC ($\gamma$), access ($\lambda$) and use ($\xi$) against true observed values from household surveys and compared against benchmark local estimates from the Bhatt-BV model. The MITN model outperformed the Bhatt-BV model for all three coverage metrics with a cluster level RMSE of 0.094, 0.145 and 0.143 for NPC, access and use respectively (see Figure 3). We also test each models' ability to reconstruct survey observations and national household aggregates. This is done by using sampled output rasters from both the Bhatt-BV and SNF models to simulate cluster level survey observations, which are subsequently aggregated into national level aggregates of NPC, access and use, and compared against estimates calculated from true survey observations using the same method. In all three metrics, the MITN significantly outperforms the Bhatt-BV model in terms of model fit with lower RMSE values. An examination of the residuals also show smaller bias in the MITN fits with exception of the access metric. The robustness of the model was also tested using a 2-fold cross validation where half of geolocated data is randomly omitted during training. The reduced data model showed similar results with only marginal increases in RMSE indicating a robust model fit.

Net attrition half-life estimates are also used for model validation and to isolate regions where model changes and new data has resulted in updated attrition estimates. Half-life estimates for the MITN were compared against those published by Bertozzi-Villa et al. in 2021 and shown in Figure 4. The MITN model covers a total of 44 countries, an increase of 4 countries (Eswatini, Sao Tome and Principe, Namibia and Botswana) from the original Bhatt-BV analysis. Attrition half-lives between both models are consistent for a majority of countries. However, the MITN tends to produce more extreme half-life estimates where countries with Bhatt-BV half-lives below 1.5 (Mauritania, Benin, Gambia) have lower MITN estimates, with the reverse being observed for those with Bhatt-BV half-lives above 3 (Congo, Cameroon). With the exception of Mauritania, Benin, Mali, Guinea-Bissau and Cameroon where there are

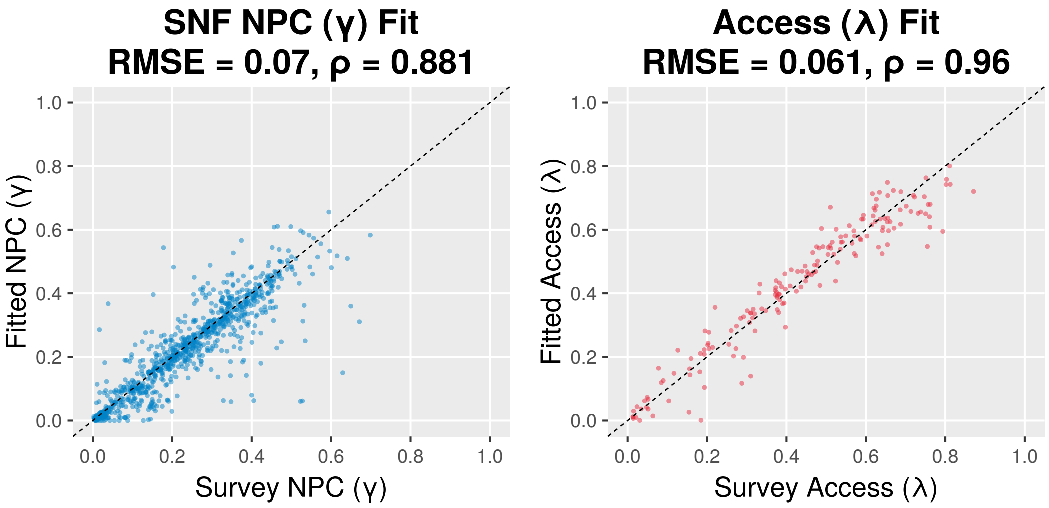


Figure 2: Fitted vs observed values for SNF and access model components at the national aggregate level. Errors are calculated against national aggregate estimates from household surveys and survey report summaries.

identifiability problems due to a lack of surveys or large gaps of missing distribution data that require imputation, a majority of analysed countries have overlaps in the the credible intervals for half-lives between both models.

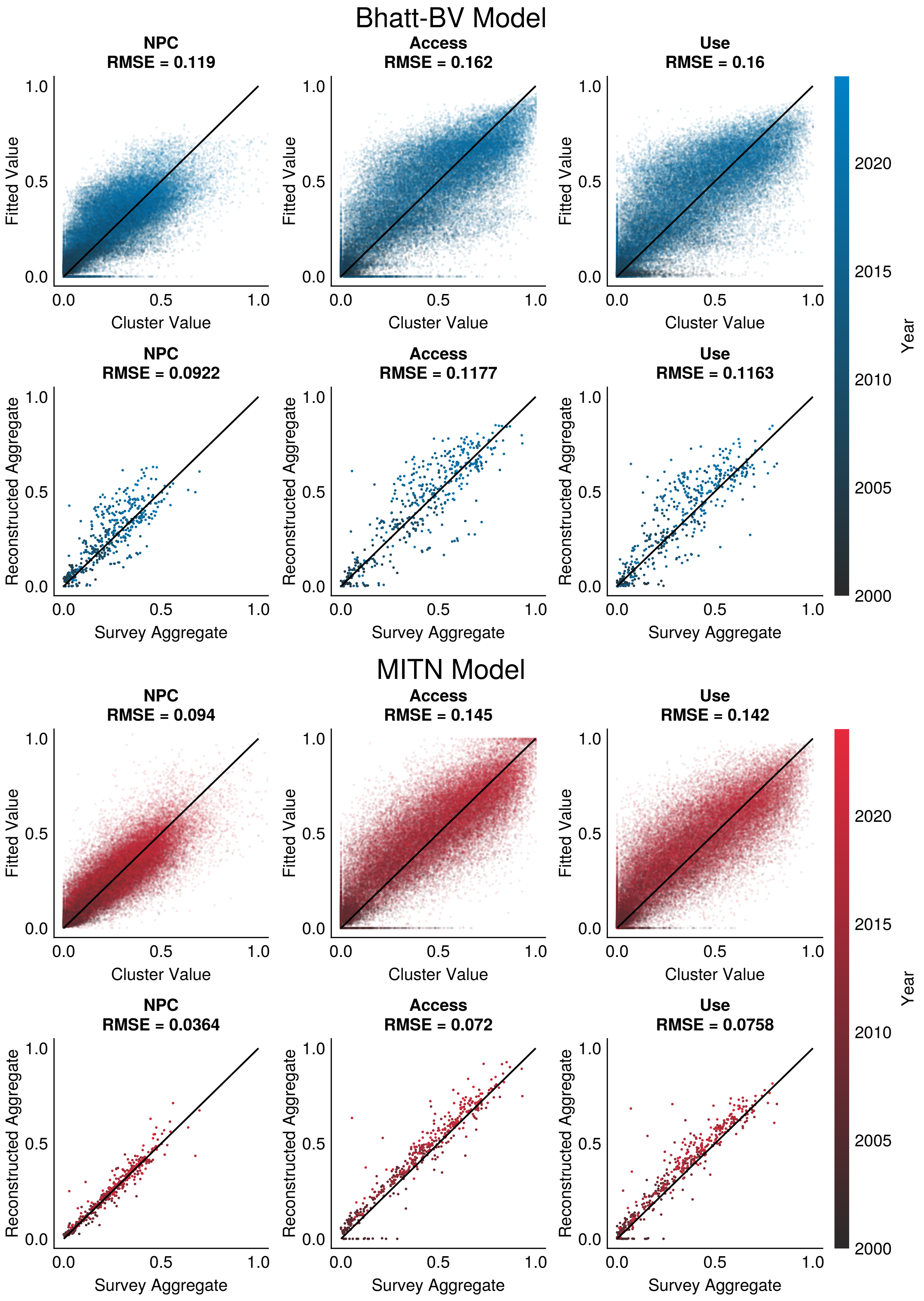


Figure 3: Fitted vs. observed values of local NPC, access and use based in the regressed spatiotemporal models. From the left to right, outputs from each model are fed into subsequent ones to construct predictions.

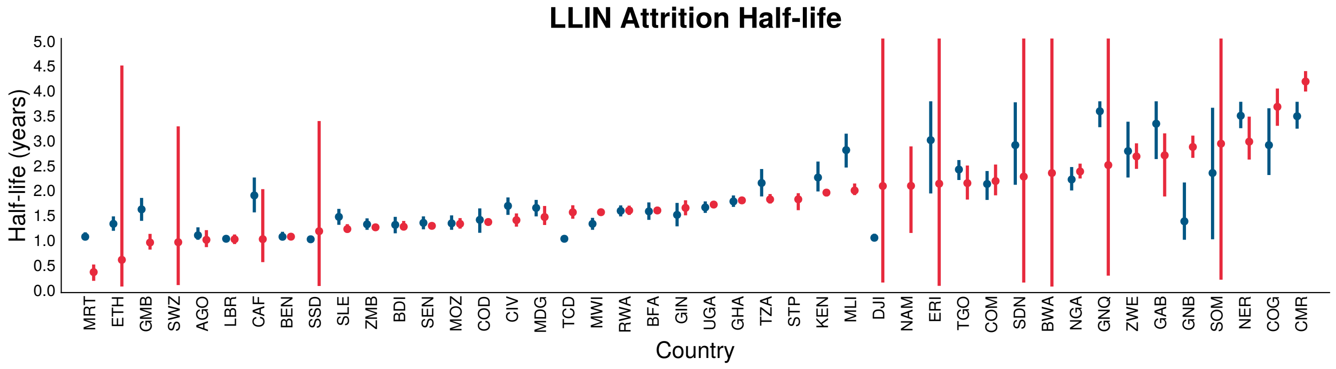


Figure 4: Estimated net attrition halflives for LLINs in analysed countries. Median and 95% credible interval show for the Bhatt-BV model (blue) and MITN model (red). Countries with large uncertainties do not have available household data and only the prior is shown.

### 6 National SNF ITN Coverage Summaries

SNF model estimates of NPC and access, related attrition curves (cITN and LLIN), and net demography with respect to age and type. Attrition of next generation LLINs (i.e. PBO, DAI) are assumed to have the same attrition parameters as LLINs due to lack of type specific survey data. Observed NPC and estimated true access values from surveys are shown as dots in the coverage metric time series. Blue vertical bars correspond to $\pm 2\sigma(t)$ (i.e. the twice the inflated uncertainty interval centred on the survey aggregate observed value). Shaded bands indicate 95% credible intervals.

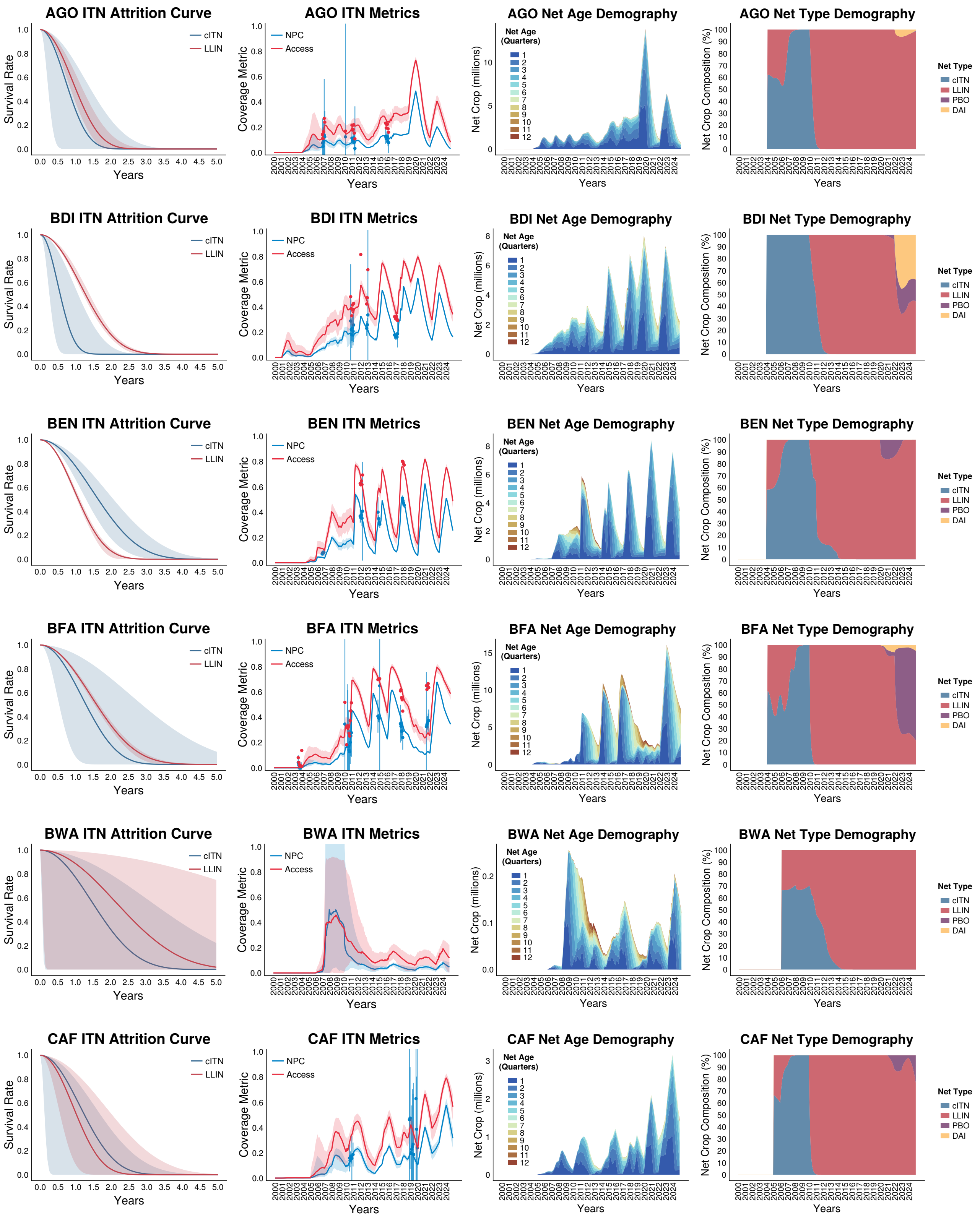

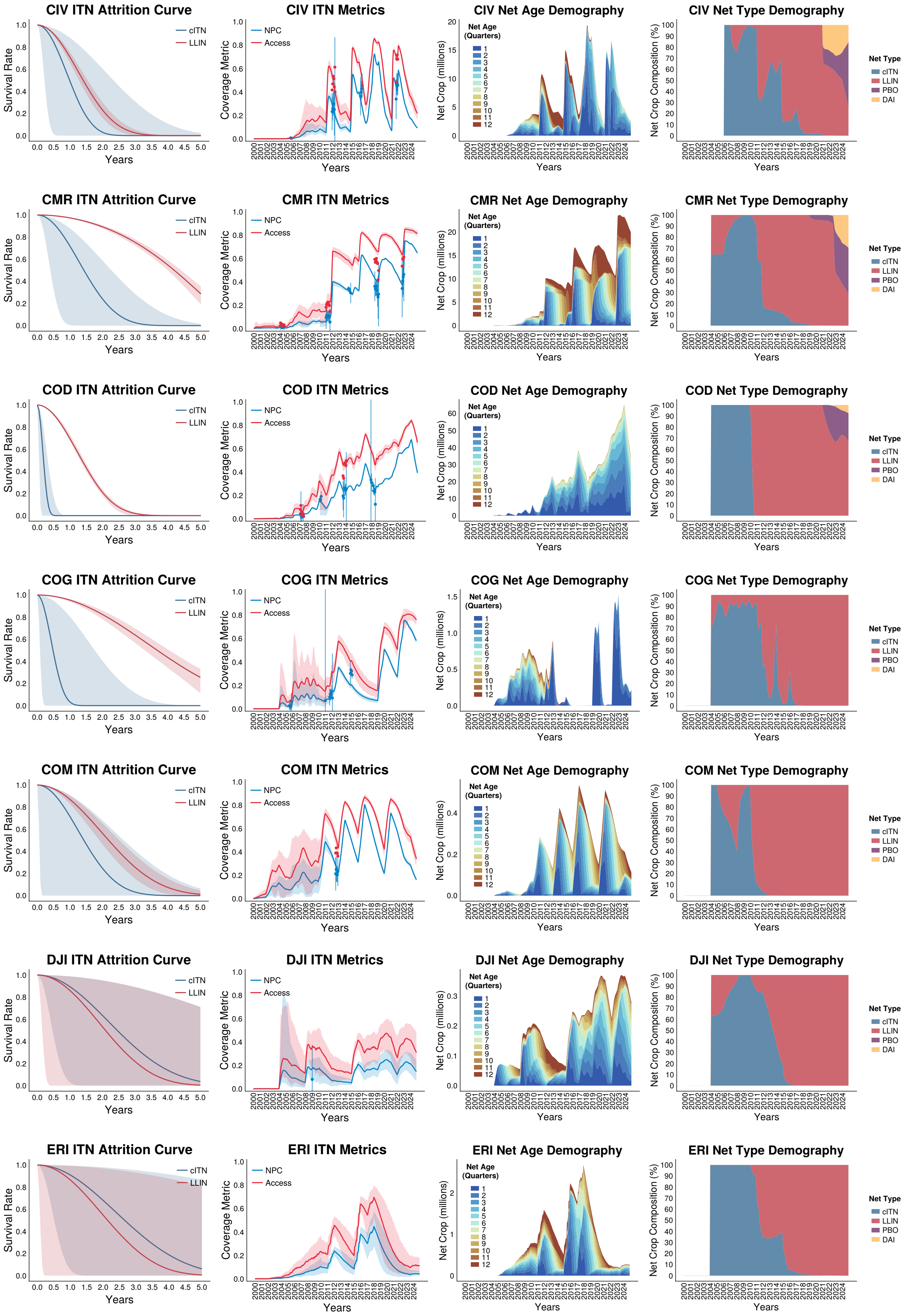
CIV ITN Attrition Curve
CIV ITN Metrics
CIV Net Age Demography
CIV Net Type Demography
CMR ITN Attrition Curve
CMR ITN Metrics
CMR Net Age Demography
CMR Net Type Demography
COD ITN Attrition Curve
COD ITN Metrics
COD Net Age Demography
COD Net Type Demography
COG ITN Attrition Curve
COG ITN Metrics
COG Net Age Demography
COG Net Type Demography
COM ITN Attrition Curve
COM ITN Metrics
COM Net Age Demography
COM Net Type Demography
DJI ITN Attrition Curve
DJI ITN Metrics
DJI Net Age Demography
DJI Net Type Demography
ERI ITN Attrition Curve
ERI ITN Metrics
ERI Net Age Demography
ERI Net Type Demography
Survival Rate
Years
cITN
LLIN
Coverage Metric
NPC
Access
Net Crop (millions)
Net Age (Quarters)
1
2
3
4
5
6
7
8
9
10
11
12
Net Crop Composition (%)
Net Type
cITN
LLIN
PBO
DAI

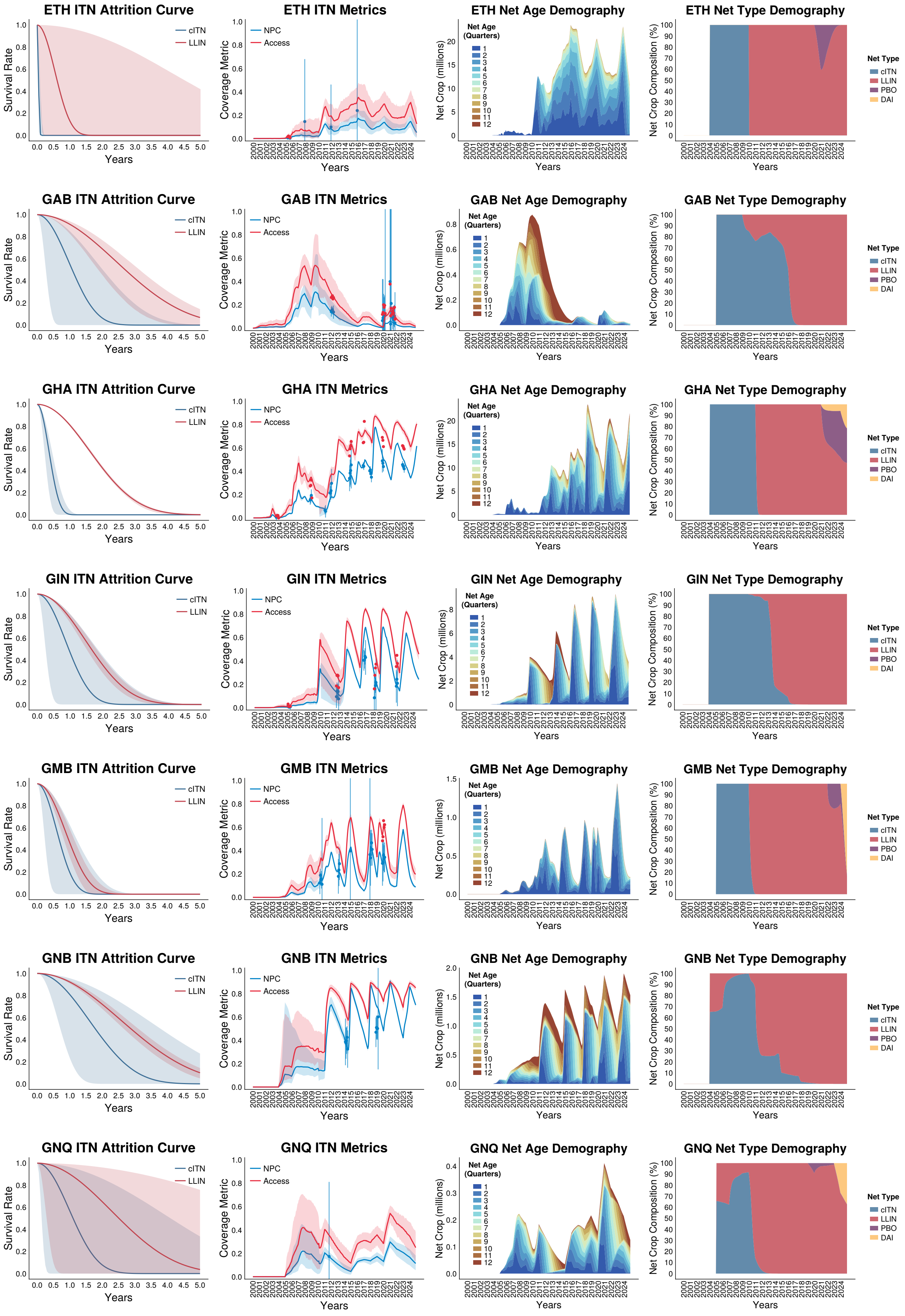
ETH ITN Attrition Curve
ETH ITN Metrics
ETH Net Age Demography
ETH Net Type Demography
GAB ITN Attrition Curve
GAB ITN Metrics
GAB Net Age Demography
GAB Net Type Demography
GHA ITN Attrition Curve
GHA ITN Metrics
GHA Net Age Demography
GHA Net Type Demography
GIN ITN Attrition Curve
GIN ITN Metrics
GIN Net Age Demography
GIN Net Type Demography
GMB ITN Attrition Curve
GMB ITN Metrics
GMB Net Age Demography
GMB Net Type Demography
GNB ITN Attrition Curve
GNB ITN Metrics
GNB Net Age Demography
GNB Net Type Demography
GNQ ITN Attrition Curve
GNQ ITN Metrics
GNQ Net Age Demography
GNQ Net Type Demography
Survival Rate
Coverage Metric
Net Crop (millions)
Net Crop Composition (%)
Years
cITN
LLIN
NPC
Access
Net Age (Quarters)
Net Type
PBO
DAI

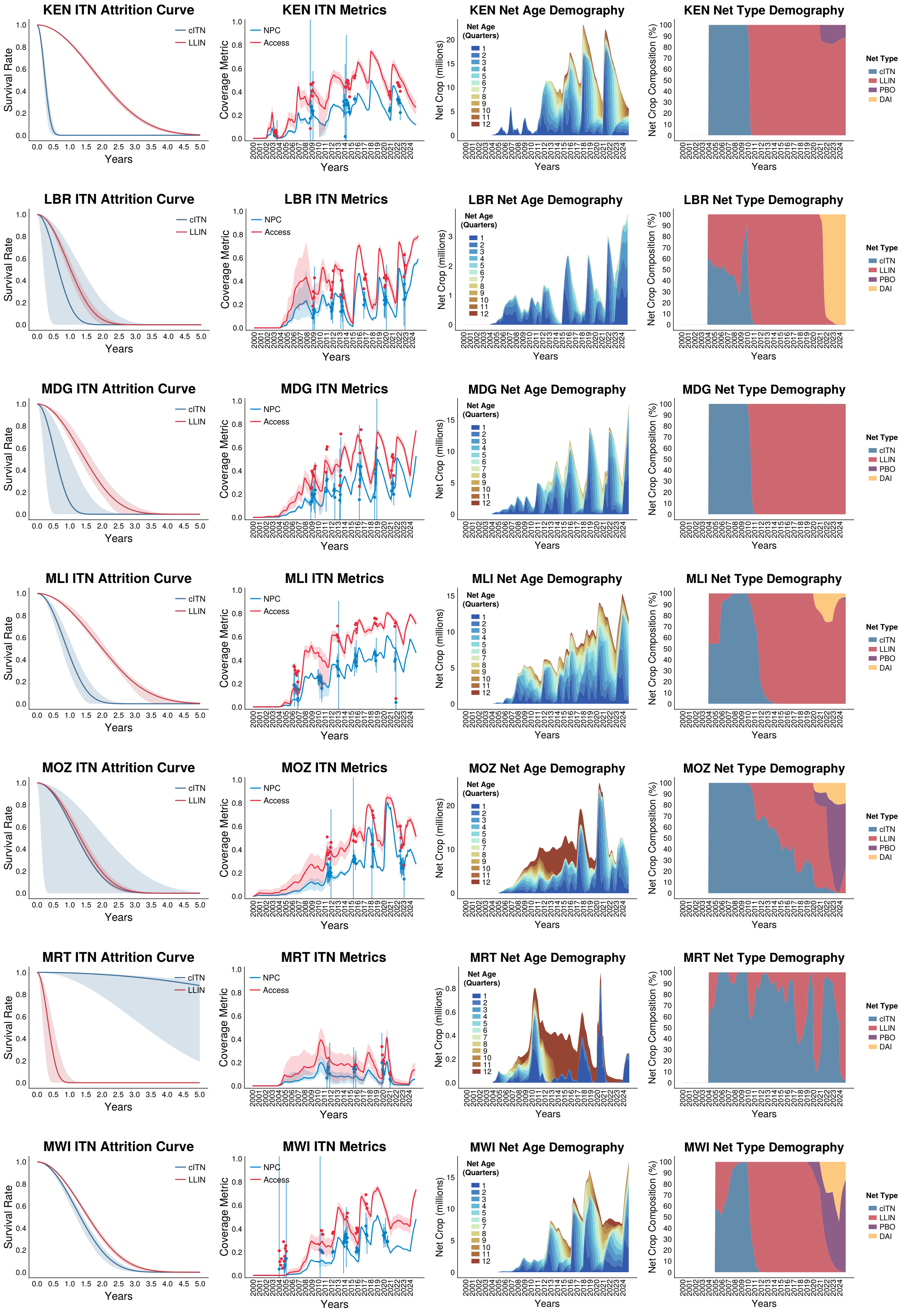
KEN ITN Attrition Curve
KEN ITN Metrics
KEN Net Age Demography
KEN Net Type Demography
LBR ITN Attrition Curve
LBR ITN Metrics
LBR Net Age Demography
LBR Net Type Demography
MDG ITN Attrition Curve
MDG ITN Metrics
MDG Net Age Demography
MDG Net Type Demography
MLI ITN Attrition Curve
MLI ITN Metrics
MLI Net Age Demography
MLI Net Type Demography
MOZ ITN Attrition Curve
MOZ ITN Metrics
MOZ Net Age Demography
MOZ Net Type Demography
MRT ITN Attrition Curve
MRT ITN Metrics
MRT Net Age Demography
MRT Net Type Demography
MWI ITN Attrition Curve
MWI ITN Metrics
MWI Net Age Demography
MWI Net Type Demography
Survival Rate
Years
cITN
LLIN
Coverage Metric
NPC
Access
Net Crop (millions)
Net Age (Quarters)
Net Crop Composition (%)
Net Type
PBO
DAI

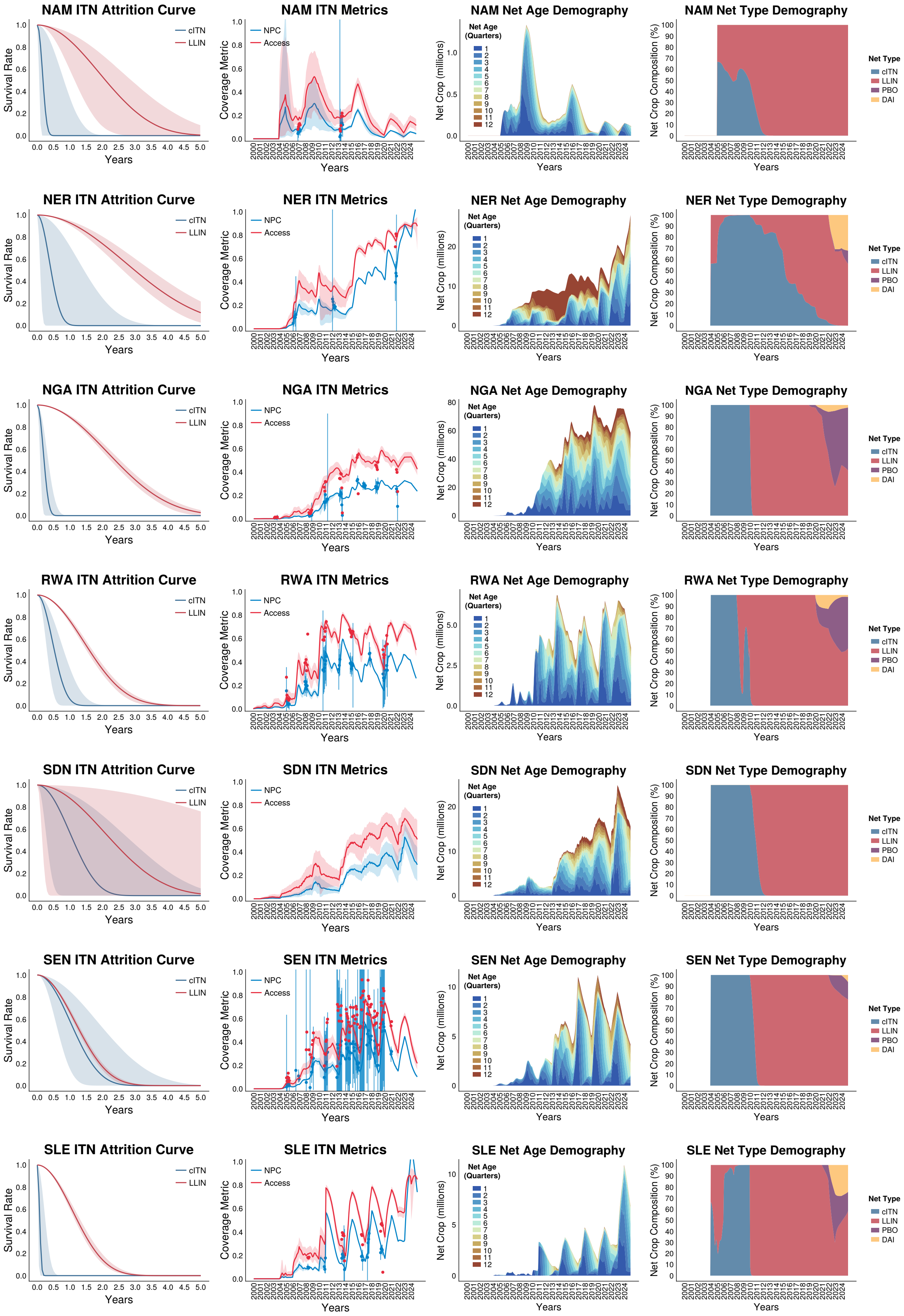
NAM ITN Attrition Curve
NAM ITN Metrics
NAM Net Age Demography
NAM Net Type Demography
NER ITN Attrition Curve
NER ITN Metrics
NER Net Age Demography
NER Net Type Demography
NGA ITN Attrition Curve
NGA ITN Metrics
NGA Net Age Demography
NGA Net Type Demography
RWA ITN Attrition Curve
RWA ITN Metrics
RWA Net Age Demography
RWA Net Type Demography
SDN ITN Attrition Curve
SDN ITN Metrics
SDN Net Age Demography
SDN Net Type Demography
SEN ITN Attrition Curve
SEN ITN Metrics
SEN Net Age Demography
SEN Net Type Demography
SLE ITN Attrition Curve
SLE ITN Metrics
SLE Net Age Demography
SLE Net Type Demography
Survival Rate
Coverage Metric
Net Crop (millions)
Net Crop Composition (%)
Years
cITN
LLIN
NPC
Access
Net Age (Quarters)
Net Type
PBO
DAI

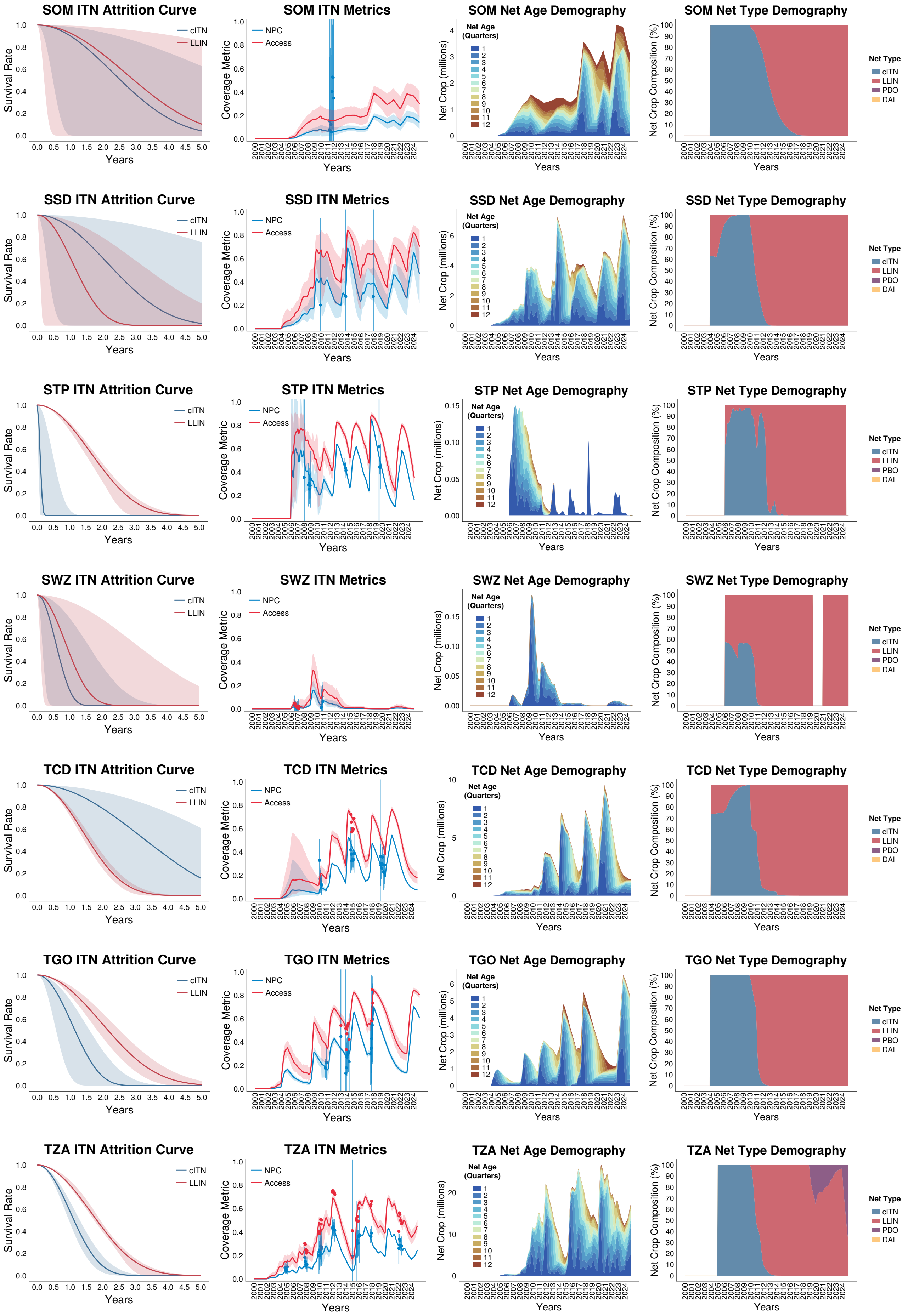
SOM ITN Attrition Curve
Survival Rate
Years
cITN
LLIN
SOM ITN Metrics
Coverage Metric
NPC
Access
SOM Net Age Demography
Net Crop (millions)
Net Age (Quarters)
SOM Net Type Demography
Net Crop Composition (%)
Net Type
cITN
LLIN
PBO
DAI
SSD ITN Attrition Curve
SSD ITN Metrics
SSD Net Age Demography
SSD Net Type Demography
STP ITN Attrition Curve
STP ITN Metrics
STP Net Age Demography
STP Net Type Demography
SWZ ITN Attrition Curve
SWZ ITN Metrics
SWZ Net Age Demography
SWZ Net Type Demography
TCD ITN Attrition Curve
TCD ITN Metrics
TCD Net Age Demography
TCD Net Type Demography
TGO ITN Attrition Curve
TGO ITN Metrics
TGO Net Age Demography
TGO Net Type Demography
TZA ITN Attrition Curve
TZA ITN Metrics
TZA Net Age Demography
TZA Net Type Demography

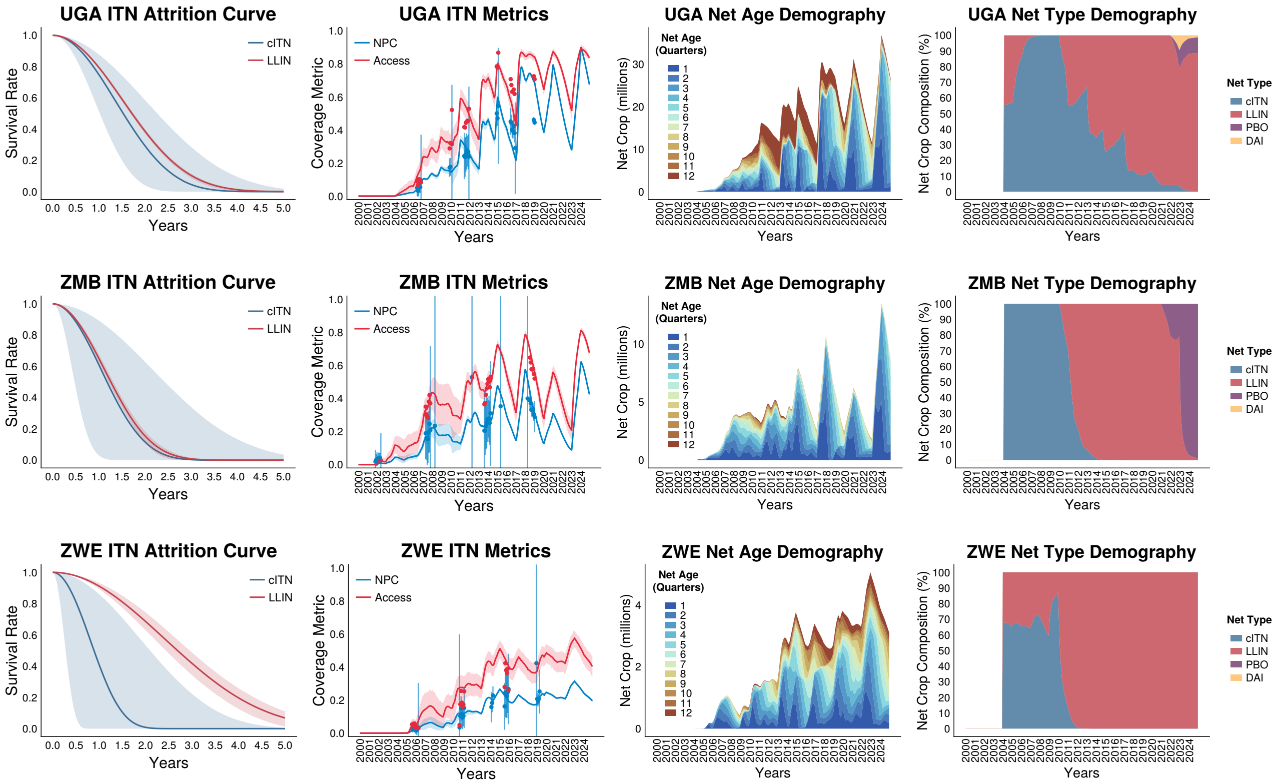
UGA ITN Attrition Curve
Survival Rate
Years
cITN
LLIN
UGA ITN Metrics
Coverage Metric
NPC
Access
UGA Net Age Demography
Net Crop (millions)
Net Age (Quarters)
UGA Net Type Demography
Net Crop Composition (%)
Net Type
cITN
LLIN
PBO
DAI
ZMB ITN Attrition Curve
ZMB ITN Metrics
ZMB Net Age Demography
ZMB Net Type Demography
ZWE ITN Attrition Curve
ZWE ITN Metrics
ZWE Net Age Demography
ZWE Net Type Demography

### 7 ITN Coverage Maps 2005-2024

#### 7.1 Coverage Maps

Spatiotemporal maps of final ITN coverage metrics (mean net age, NPC, access, use and utilisation) averaged annually and shown for the years 2000-2024.

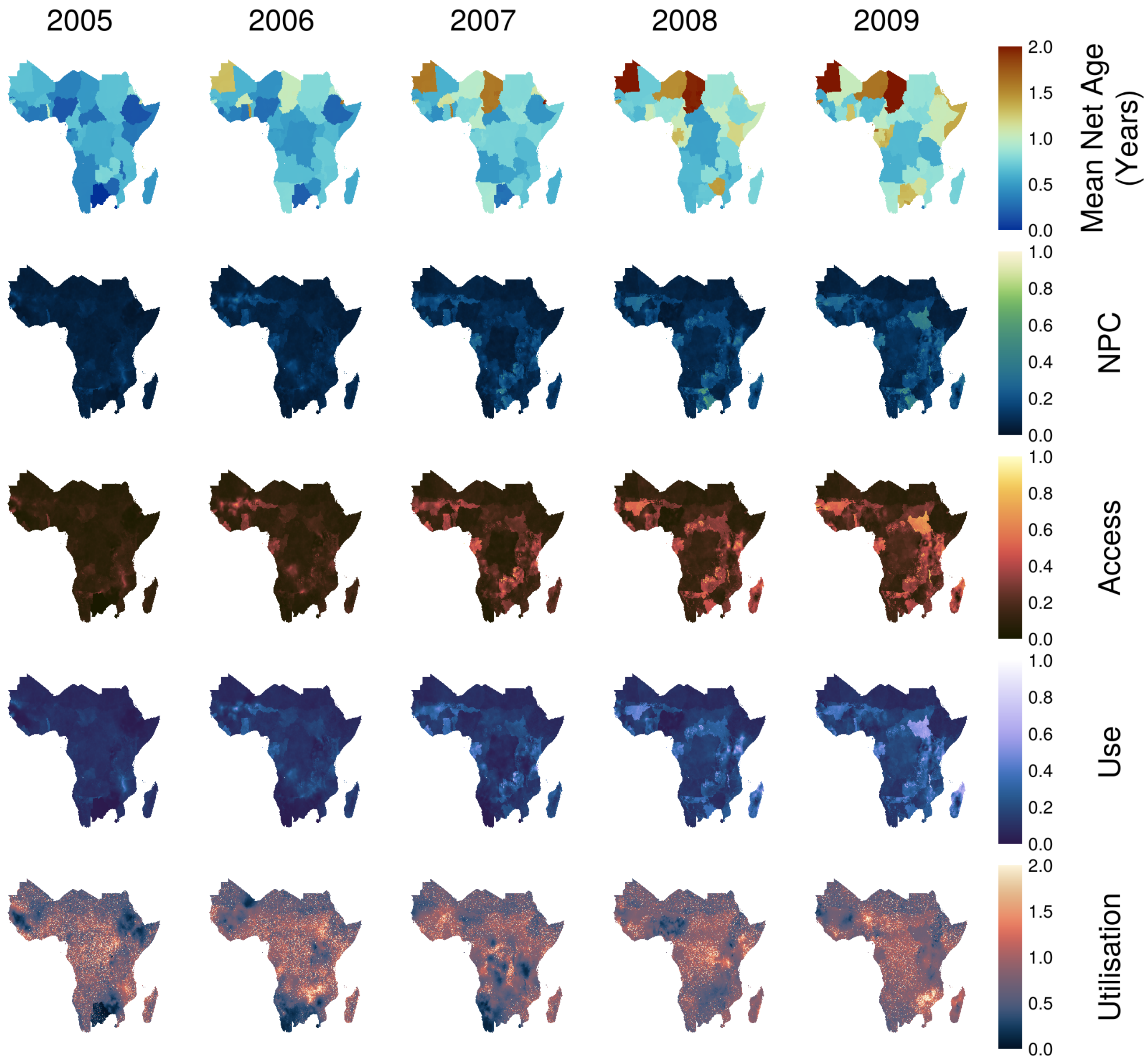

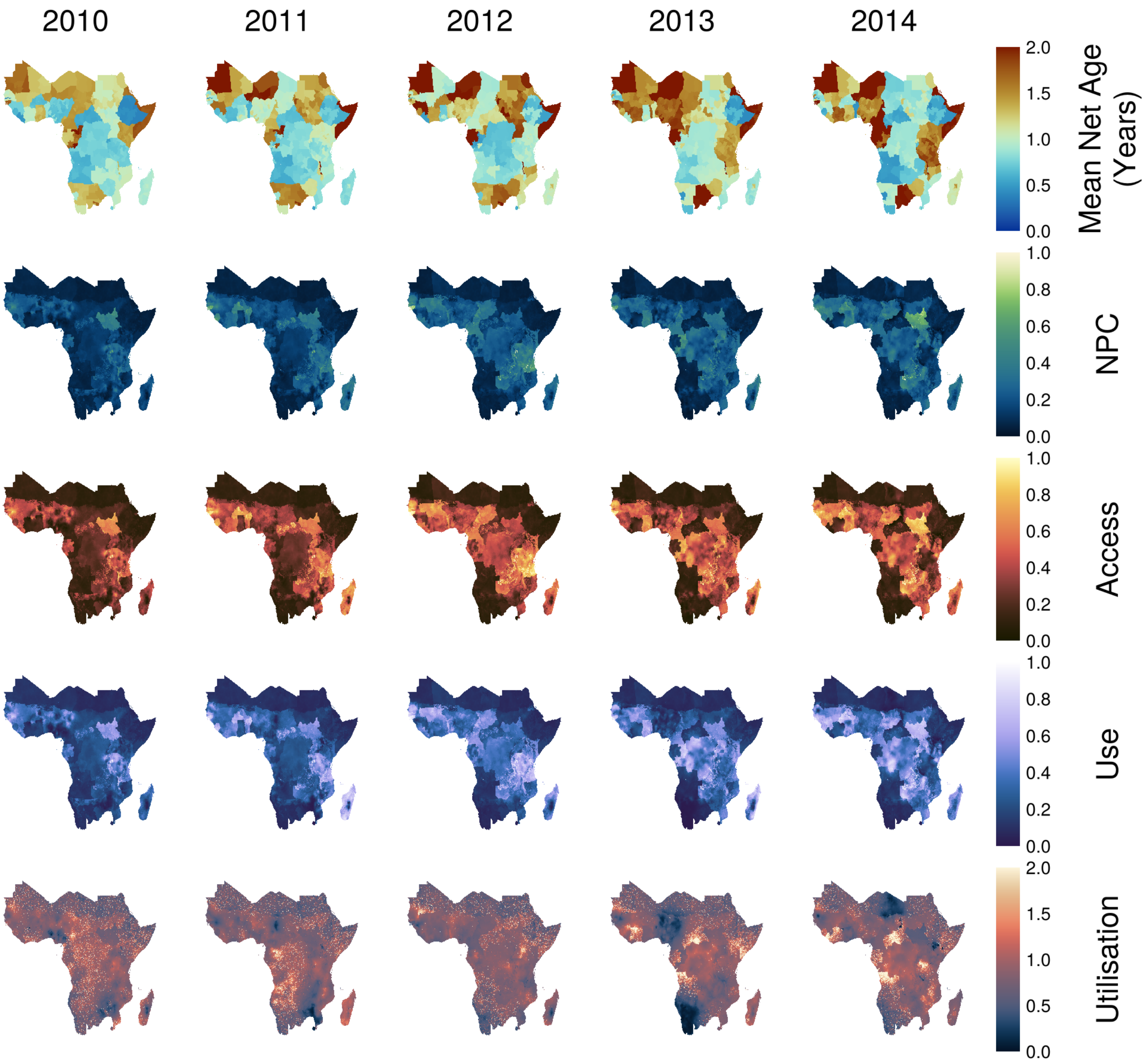

2010
2011
2012
2013
2014
2.0
1.5
1.0
0.5
0.0
Mean Net Age (Years)
1.0
0.8
0.6
0.4
0.2
0.0
NPC
1.0
0.8
0.6
0.4
0.2
0.0
Access
1.0
0.8
0.6
0.4
0.2
0.0
Use
2.0
1.5
1.0
0.5
0.0
Utilisation

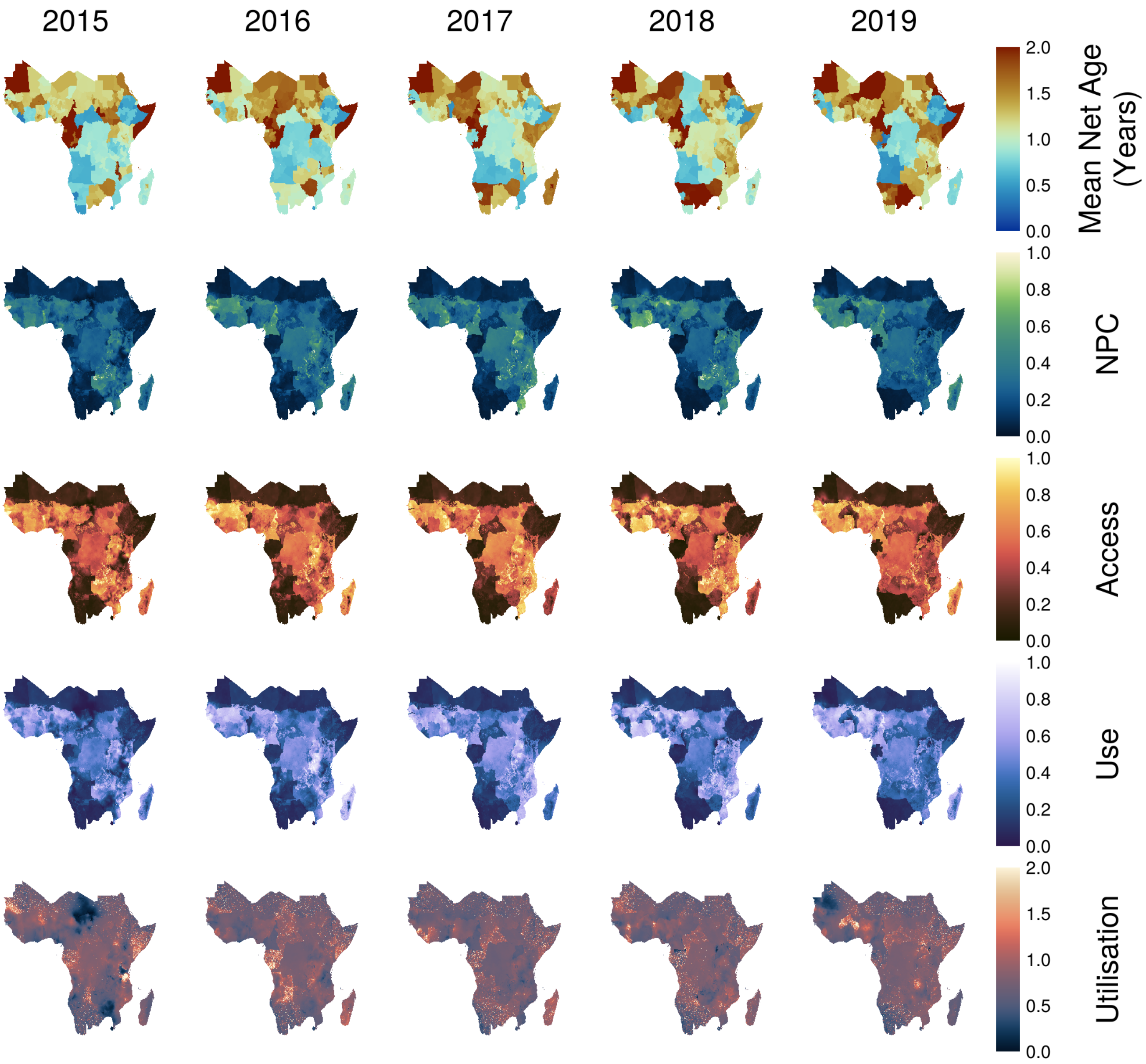
2015
2016
2017
2018
2019
2.0
1.5
1.0
0.5
0.0
Mean Net Age (Years)
1.0
0.8
0.6
0.4
0.2
0.0
NPC
1.0
0.8
0.6
0.4
0.2
0.0
Access
1.0
0.8
0.6
0.4
0.2
0.0
Use
2.0
1.5
1.0
0.5
0.0
Utilisation

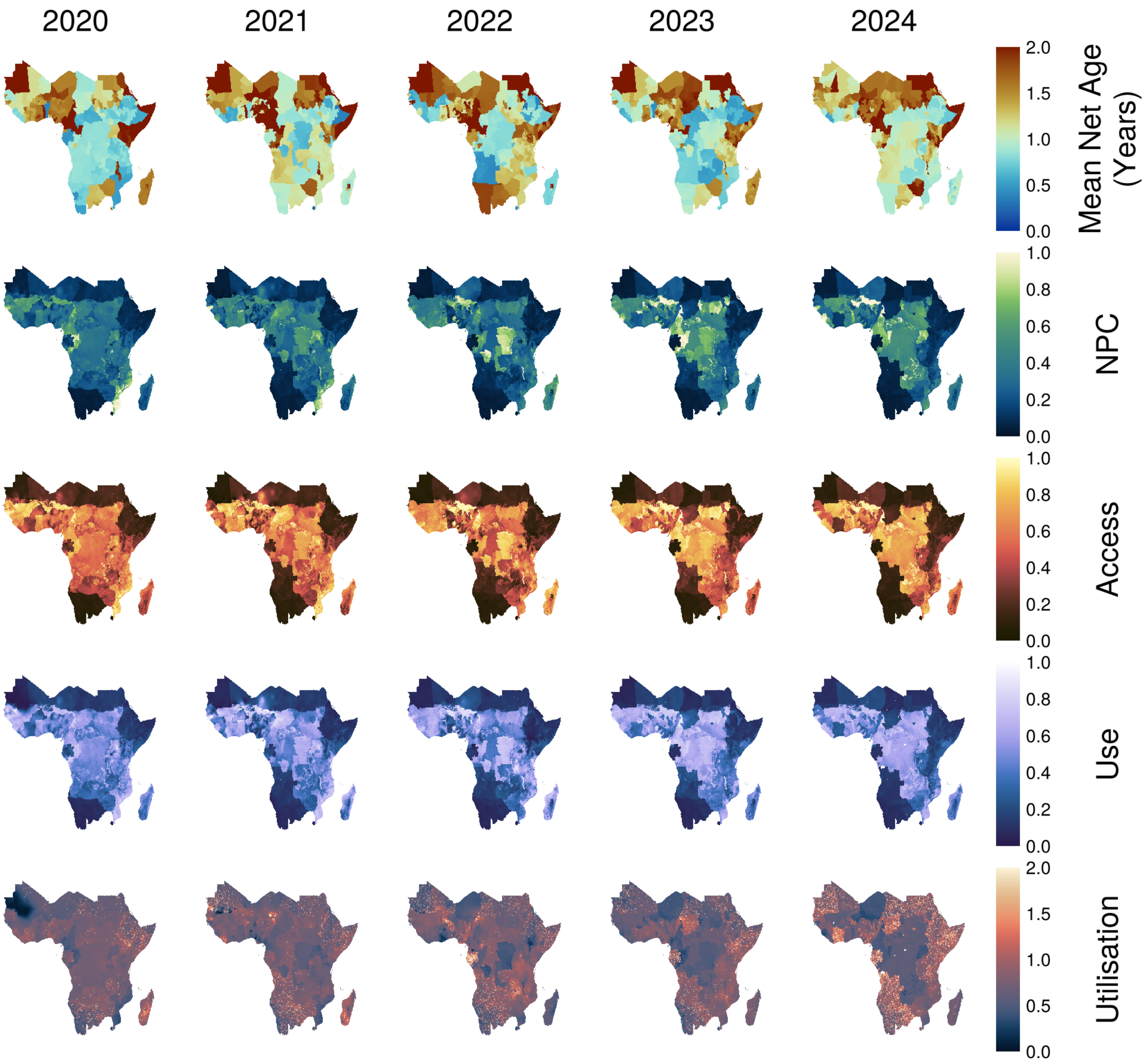
2020
2021
2022
2023
2024
2.0
1.5
1.0
0.5
0.0
Mean Net Age (Years)
1.0
0.8
0.6
0.4
0.2
0.0
NPC
1.0
0.8
0.6
0.4
0.2
0.0
Access
1.0
0.8
0.6
0.4
0.2
0.0
Use
2.0
1.5
1.0
0.5
0.0
Utilisation

#### 7.2 Uncertainty Quantification of Spatiotemporal ITN Coverage

For SNF estimates of the model, attrition and redistribution parameter posteriors can be propagated forward to calculate estimates of uncertainty for national (Admin 0) and subnational (Admin 1) aggregates of ITN coverage excluding spatial variability. These uncertainties can be propagated forward into the spatiotemporal model to produce posterior draws of rasters. Raster samples can be averaged to produce mean rasters for each month (or aggregated in to years) such as those in Section (SECION REF). However, unlike outputs of the SNF, the definition of uncertainty for rasters is non-trivial.

Let $\mathcal{M} \in \mathbb{R}^2 \times T \times \mathbb{R}$ be a raster where the three components $\mathbb{R}^2$, $T = \mathbb{R}^+$ and $\mathbb{R}$ represent the space, time and raster value. Alternatively, a raster can also be represented as a scalar function where

$$\mathcal{M} : \mathbb{R}^2 \times T \to \mathbb{R}.$$

In the case of the deviation rasters (represented with hat notation) output by the MITN model, they can be written as

$$\hat{\psi}(x,t) = \underbrace{\sum_i \beta_i X_i(x,t)}_{\text{spatiotemporal covariates}} + \underbrace{F_{\hat{\psi}}(x,t)}_{\text{random field}}.$$

Assuming that estimates of covariates $X_i$ are reliable with small uncertainty, all uncertainty in the raster $\hat{\psi}$ can be attributed to the random field $F_{\hat{\psi}}$. For subsequent parts of this section, we will refer to $\mathcal{M}(x,t)$ as any raster that describes a coverage quantity of interest, whose posterior follows some distribution $\mathcal{P}_{\mathcal{M}}$ defined over the space of all possible rasters ($\mathbb{R}^2 \times \mathbb{R}^+ \times \mathbb{R}$). Therefore, we seek to quantify the uncertainty associated with the random raster (variable) $\mathcal{M}$.

One can attempt to directly calculate the uncertainty of $\mathcal{M}$ across its entire ($\mathbb{R}^2 \times \mathbb{R}^+ \times \mathbb{R}$) domain. This retrieves the full uncertainty structure of $\mathcal{P}_{\mathcal{M}}$, but is both computationally expensive with little use in practice in the context of policy decisions and quantifying ITN coverage uncertainties. Instead, one can define a sample summary statistic $S(\mathcal{M}) : \mathbb{R}^2 \times \mathbb{R}^+ \times \mathbb{R} \to \mathbb{R}$ whose uncertainty can be taken as representative proxy of the true uncertainty provided $S$ measures a quantity of interest. Consequently, we have

$$S(\mathcal{M}) \sim \mathcal{P}^*_{\mathcal{M}},$$

where $\mathcal{P}^*_{\mathcal{M}}$ is the univariate distribution of the sample statistic $S$ calculated over randomly drawn posterior sample rasters $\mathcal{M}$. A reasonable choice for $S(\mathcal{M})$ is

$$S(\mathcal{M}; \mathcal{D}) = \int_{\mathcal{D}} \mathcal{M}(x,t)\, d\mathcal{D},$$

where $\mathcal{D}$ is a chosen domain of interest over which the statistic is calculated. Here, we make the simplification to keep the temporal component constant and only define $\mathcal{D}$ as a spatial domain.

Because $S$ is dependent on the domain $\mathcal{D}$, a choice needs to be made on its size and location which can vary between two extremes: - Small spatial scale: Set $\mathcal{D}$ to be equal to the spatial resolution of the raster (i.e. one pixel). This would be equivalent to calculating the marginal uncertainties of each pixel. - Large spatial scale: Let $\mathcal{D}$ to be equal to the union of the entire domain of the raster (i.e. the continent). Both extreme choices contain several key drawbacks. The first approach makes the assumption that uncertainties between neighbouring regions (e.g. pixels) are independent. However, because the random effects are represented as a collection of spatial Gaussian processes with some underlying covariance structure, close neighbours are generally not independent. Pixel-level marginals will result in overestimated values of uncertainty. The opposite problem is true for the latter case of large spatial scales where much of the spatial variation in uncertainty is lost.

Instead, we argue that $\mathcal{D}$ should be chosen to be of an intermediate spatial scale with boundaries determined such that for any two neighbouring regions $\mathcal{D}_i$ and $\mathcal{D}_j$, one can reasonably assume independence $\mathcal{P}^*_{\mathcal{M}}(\mathcal{D}_i) \perp \mathcal{P}^*_{\mathcal{M}}(\mathcal{D}_j)$. Ideally the spatial scale of $\mathcal{D}$ should be larger than the spatial scale of the random Gaussian processes random field $F$.

Reasonable choices of national or subnational boundaries aims to provide uncertainty estimates for quantities that are most likely to be utilised by stakeholders, leverages the mathematical capabilities of the model, whilst providing a more nuanced and representative estimate of uncertainty. For illustration, a comparison of the uncertainty calculated using the pixel level marginal (small spatial scale) and subnational region boundaries (intermediate spatial scales) are presented in Figure 5. Estimates for mean and uncertainty values for the entire historical period (2000-2024) across all countries and subnational regions can be provided upon direct request.

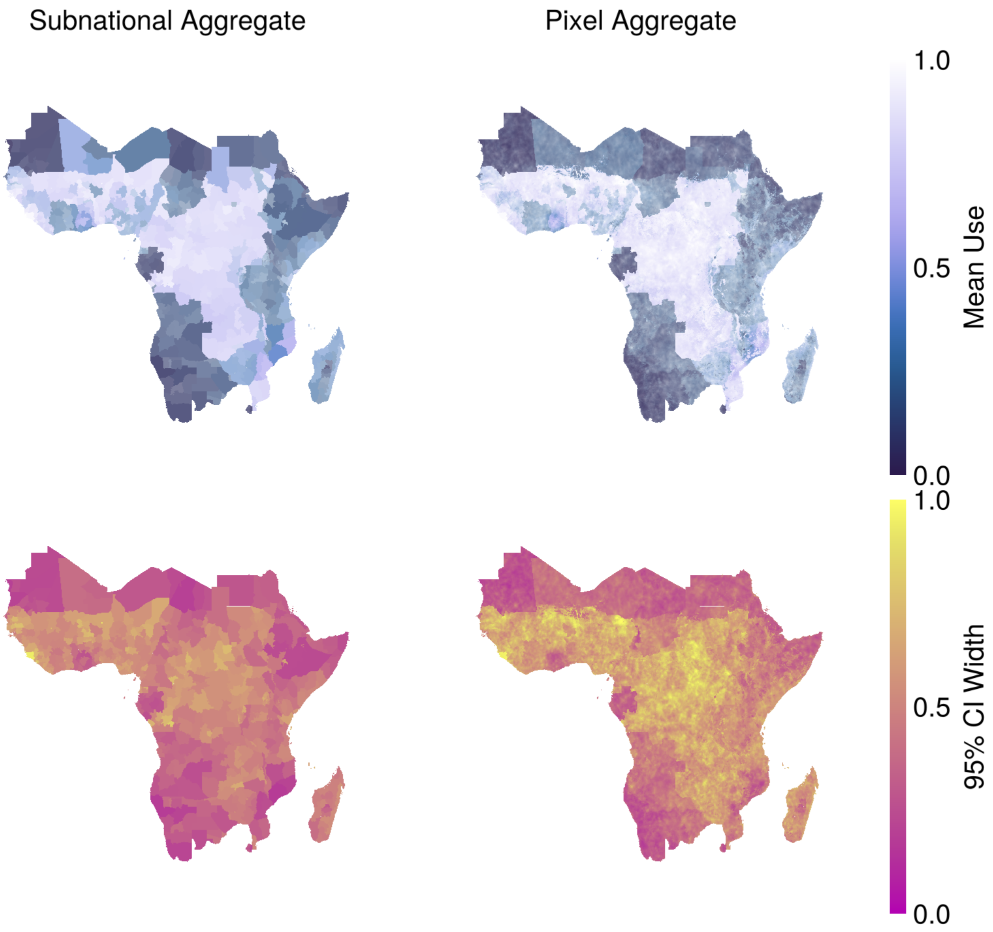


Figure 5: Map of mean ITN Use with transparency weighted by the width of the 95% credible interval (top), and maps of the 95% credible interval width (bottom) calculated for subnational aggregates and pixel level estimates. The latter approach calculated with pixel-level marginals produces higher levels of uncertainty, whilst the subnational aggregates have a tighter bound.

### 8 Country ITN Utilisation and Use Rates

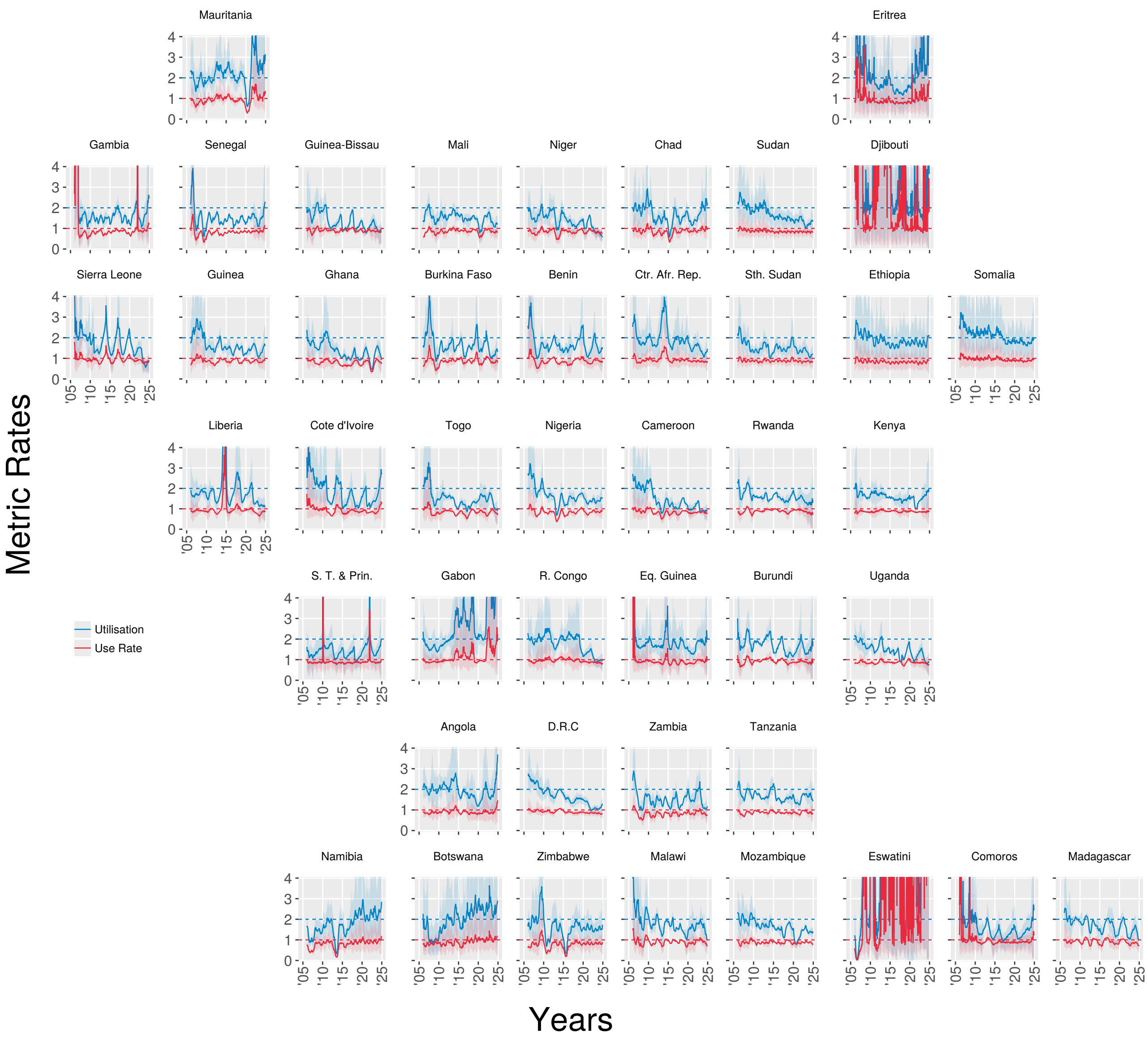


Figure 6: Utilisation and use rate time series extracted from produced MITN coverage raster maps. Shaded bands indicate 95% credible intervals.

### 9 Macroeconomic Trends

In the absence of alternative high-efficacy and low-cost interventions, growing populations in the African region are expected to increase the need and thus demand of ITNs. To quantify this, we track the annual percentage change in the 2 year moving average of net crop and population across all 44 modelled countries in the African regions (see Figure (7)). Fuelled by the growth in net crop exceeded population growth across Africa until 2019, from 2019 to 2022, growth in these factors was approximately equal, and since 2022 population growth has outpaced net crop growth. A small uptick in net crop during in the year 2023 is attributed to net distributions by several countries and it is not clear if this is part of a long term trend. Therefore, we project that current ITN distribution strategies will result in a continued period where population growth outpaces net availability.

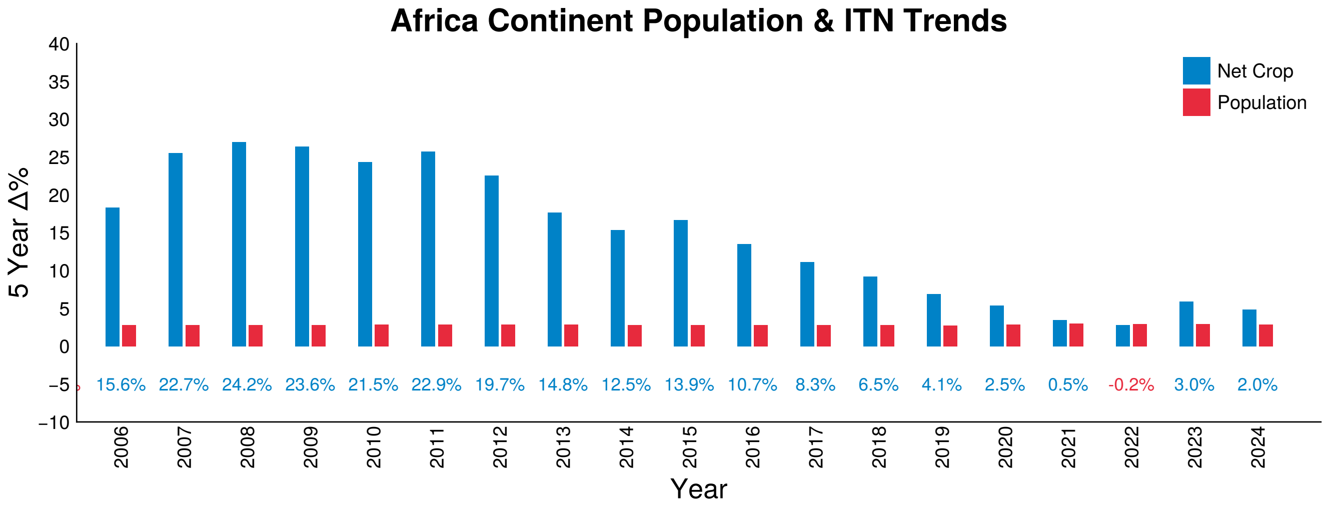


Figure 7: Figure shows annual percentage change in the 2 year moving average for total population and net crop across the 44 modelled countries in the African region. Coloured numbers indicate the difference in percentage growth between net crop (blue) and population (red).
